# Thalamic burst and tonic firing selectively indicate patients’ consciousness level and recovery

**DOI:** 10.1101/2024.09.21.24313796

**Authors:** Huan Wang, Yongxiang Hu, Qianqian Ge, Yuanyuan Dang, Yi Yang, Long Xu, Xiaoyu Xia, Peng Zhang, Sheng He, Steven Laureys, Yan Yang, Jianghong He

## Abstract

Patients with disorders of consciousness exhibit severe declines in arousal and awareness, alongside anomalous functional brain connections and aberrant neuronal activities^1–4^. Yet, the diagnostic error of patients’ consciousness states can reach up to forty percent^5,6^, resulting in a worse prognosis. Neuronal mechanisms underlying the disorders are indispensable for identifying objective and intrinsic markers of consciousness. As the principal relay station between the brainstem arousal nuclei and the cerebral cortex, the thalamus has been empirically inferred to maintain consciousness and wakefulness within the brain connectome^7–12^. Here, we investigated thalamic spiking, brain connections, consciousness states, and outcomes following deep brain stimulation in 29 patients. Our study reveals that thalamic activities can signal their consciousness states. Patients diagnosed with vegetative state/unresponsive wakefulness syndrome exhibited less active neurons with longer and more variable burst discharges compared to those in a minimally conscious state. Furthermore, as a direct deep brain stimulation site, neuronal profiles in the centromedian/parafascicular complex of the thalamus indicated whether electrostimulation here improved outcomes. Stronger tonic firing was associated with enhanced thalamocortical connections and better recovery outcomes in patients. These findings suggest that thalamic spiking signatures, including single-neuron burst discharge and tonic firing, selectively indicate the representation and alteration of consciousness. These findings provide direct neuronal and clinical evidence for understanding of thalamic contributions to disorders of consciousness.

## Main

Thalamus may modulate consciousness through its spiking activity. Thalamic dual firing modes, tonic and burst, have distinct effects on thalamocortical interactions^13–19^. Their burst modes were regulated during states of unresponsiveness or unconsciousness. Nonetheless, this knowledge about neuronal correlates of consciousness, especially at the cellular level, was derived primarily from comparisons between anesthesia, natural sleep, and wakefulness in animal models (rodents^9^, cats^7^ and monkeys^10^). Comparing data from three patients with disorders of consciousness (DoC), only a few clinical cases have shown that vegetative state/unresponsive wakefulness syndrome (VS/UWS) and minimally conscious state (MCS) differ in thalamic spiking activity^20^. The lack of neurophysiological signatures analysis in DoC patients, particularly at the single-unit level, hinders our ability to understand the disorders’ neuronal mechanisms in greater depth.

As the primary hub between the brainstem arousal nuclei and the cerebral cortex, the thalamus may function as an essential node within the neuronal network responsible for regulating arousal^1,2,8,21–24^. The mesocircuit hypothesis posits that the recovery of consciousness may be influenced by frontostriatal connections, with a specific contribution from the thalamus^1,21^. Deep brain stimulation (DBS) in the thalamus has been demonstrated to improve cognitive and behavioral functions in DoC patients^1,21,25–30^. A recent study demonstrated that DBS in the central lateral nucleus can enhance executive function in 5 conscious patients with moderate-to-severe traumatic brain injury^31^. Thalamic DBS has the potential to modulate thalamocortical and corticocortical interactions, alter activity in wide-ranging frontoparietal cortices and the cingulate, and rouse animals from stable anesthesia^10,11,32,33^. Although stimulation targeting specific nodes and connections could reactivate injured arousal networks and promote the re-emergence of consciousness, the underlying neuronal mechanisms and neuronal circuits have not yet been fully elucidated. In addition, clinical evidence indicating that the effects of arousal regulation with DBS differed among DoC patients^26,34^ speaks for a personalized prognosis^35^. Further research is required to comprehend intrinsic neuronal activities and how they contribute to DoC symptoms and outcomes following DBS, particularly through direct clinical evidence on single neurons and functional connections in DoC patients.

Here, we conducted a retrospective investigation into the neuronal mechanisms underlying the representation and alteration of consciousness in 29 patients diagnosed with DoC. We employed multi-model measurements utilized in clinical treatments. Specifically, we analyzed thalamic spiking activity collected as patients recovered from anesthesia during neurosurgery. We statistically evaluated relationships between the spiking activity of thalamic neurons, preoperative resting-state functional magnetic resonance imaging (rs-fMRI) data, and clinical neurobehavioral assessments before and after DBS within a 12-month period (Extended Data Fig. 1a).

## Results

### Neuronal signatures of thalamic neurons indicate DoC patients’ consciousness levels

We first describe thalamic microelectrode recordings. All recorded single units were offline sorted and remapped according to reconstructed DBS lead trajectories (Fig. 1a, bilateral leads from patient No.26 as an example; Extended Data Fig. 1c, leads for all 29 patients). As depicted in Fig. 1b, the lead has traversed the ventral lateral nucleus (VL), and the central lateral nucleus (CL) before landing its C0 contact into the centromedian/parafascicular complex (CM/Pf) of the thalamus. Neuronal activities were recorded from a total of 682 thalamic sites in 29 patients for subsequent analyses (Fig. 1c).

**Fig. 1.**
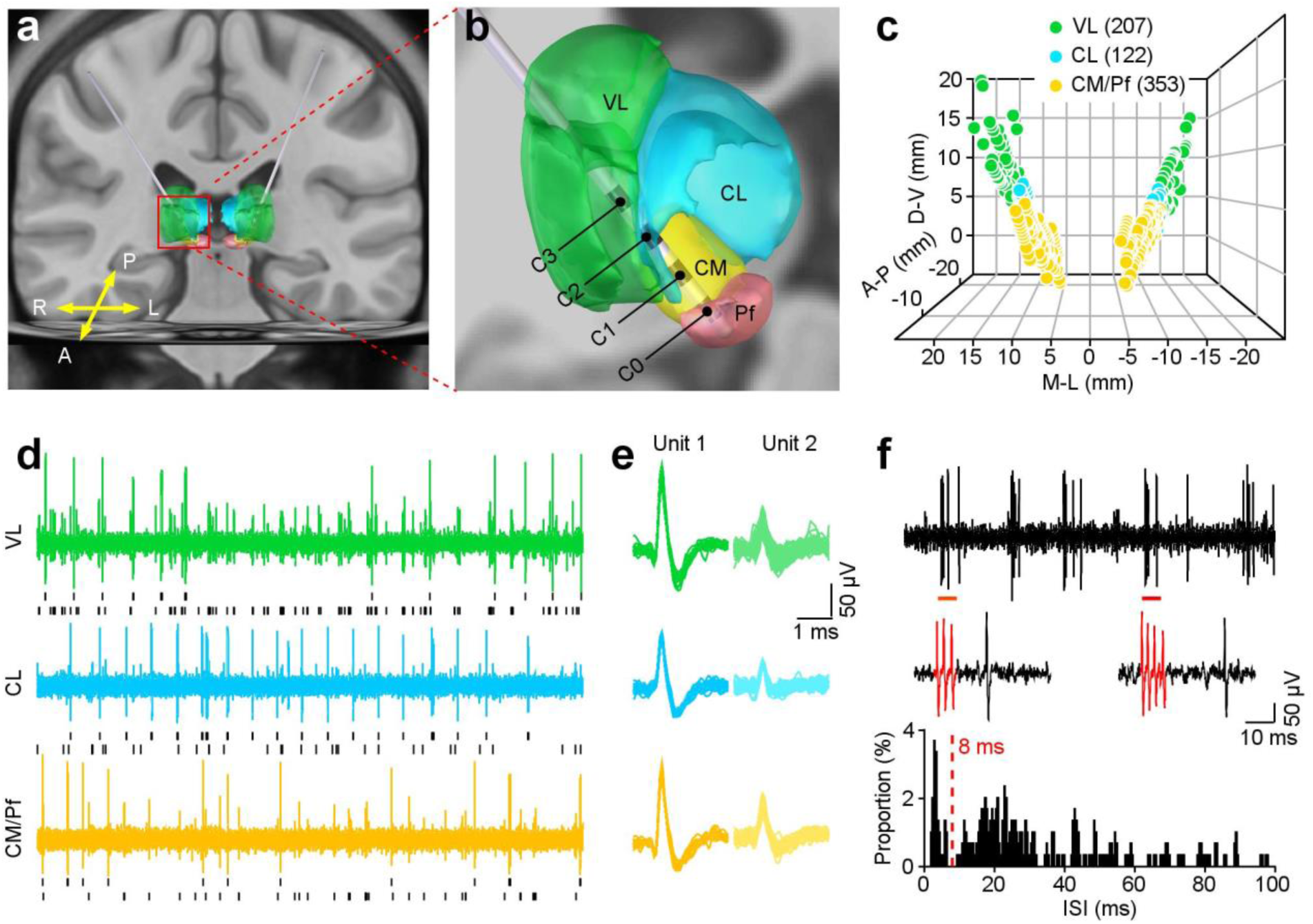
Neuronal spiking recording and DBS applications in the human thalamus. **a**, Example trajectories in a DoC patient show bilateral implantation of quadripolar DBS leads into the thalamus. A, anterior; P, posterior; R, right; L, left. **b**, Magnification of the right hemisphere (red box in **a**) shows the anatomical structures of three thalamic nuclei (green: ventral lateral nucleus (VL), blue: central lateral nucleus (CL), yellow: centromedian nucleus (CM), and pink: parafascicular nucleus (Pf)), and lead contacts’ locations. **c**, Three-dimensional positions of 682 thalamic recording sites in 29 patients. **d**, **e**, Examples of neuronal spiking activity in the thalamic nuclei from a patient. (**d**) raw spike traces for 5 seconds, with short black lines representing spike trains of sorted neurons. Their spike waveforms are shown in **e**. **f**, Burst and tonic modes of an example thalamic neuron. From top to bottom: raw spike trace of 1 second with red bars indicating two burst events; the two bursts shown with an expanded time base (red traces with 3 and 4 spikes in each one); distribution of its ISIs with 15.44% of spikes in bursts defined by an inter-spike interval of less than 8 ms.

The electrophysiological signatures of single units and multi units in the three thalamic nuclei were investigated (Figs. 1d,e). For *a single neuron*, we analyzed the single-unit frequency (SUF) and the geometric coefficient of variation (GCV) of its inter-spike interval (ISI). 94.49% of recorded thalamic neurons exhibit both tonic and burst firing modes (Fig. 1f). A burst event was identified by clusters of at least two spikes with ISIs shorter than 8 milliseconds. We categorized spikes and calculated their firing rates of burst spikes (BS) and tonic spikes (TS) based on whether or not they were in burst mode. The burst event frequency (BEF, burst events per second), the burst length (BL, spike numbers per burst), and its coefficient of variation (BL-CV) were also calculated. For *multiple neurons*, we computed the multi-unit frequency (MUF), the sample entropy (SpEn), and the unit number (UN) at each recording site. Here, ten spike-based neuronal signatures were extracted from DoC patients to characterize a panoramic view of neuronal spiking activity in the thalamus.

We employed the JFK Coma Recovery Scale-Revised (CRS-R)^36^, a validated neurobehavioral assessment designed to characterize and monitor patients’ consciousness states (see Extended Data Table 1 for details). To capture the relationship between neuronal activities in the thalamus and consciousness levels, we grouped patients according to their preoperative states of consciousness: VS/UWS (n = 17 patients) and MCS (n = 12 patients) (Fig. 2a). To evaluate neuronal signatures for both single units and multiple units at each recording site without bias towards specific types of thalamic neurons, we performed analyses on the mean responses across neurons when multiple neurons were recorded at that site. Further, we independently compared 10 neuronal signatures in the thalamic nuclei, VL, CL, and CM/Pf (Fig. 2b), and identified neuronal signatures exhibited differences between the two states of consciousness (Fig. 2c). Patients in VS/UWS with a lower level of consciousness exhibited less active thalamic neurons, characterized by longer and more variable burst discharges in the CM/Pf, and lower burst firing activities in the CL and VL (Figs. 2b,c), compared to those in MCS. These signatures in thalamic nuclei, including burst discharges of single neurons, effectively distinguish VS/UWS from MCS.

**Fig. 2.**
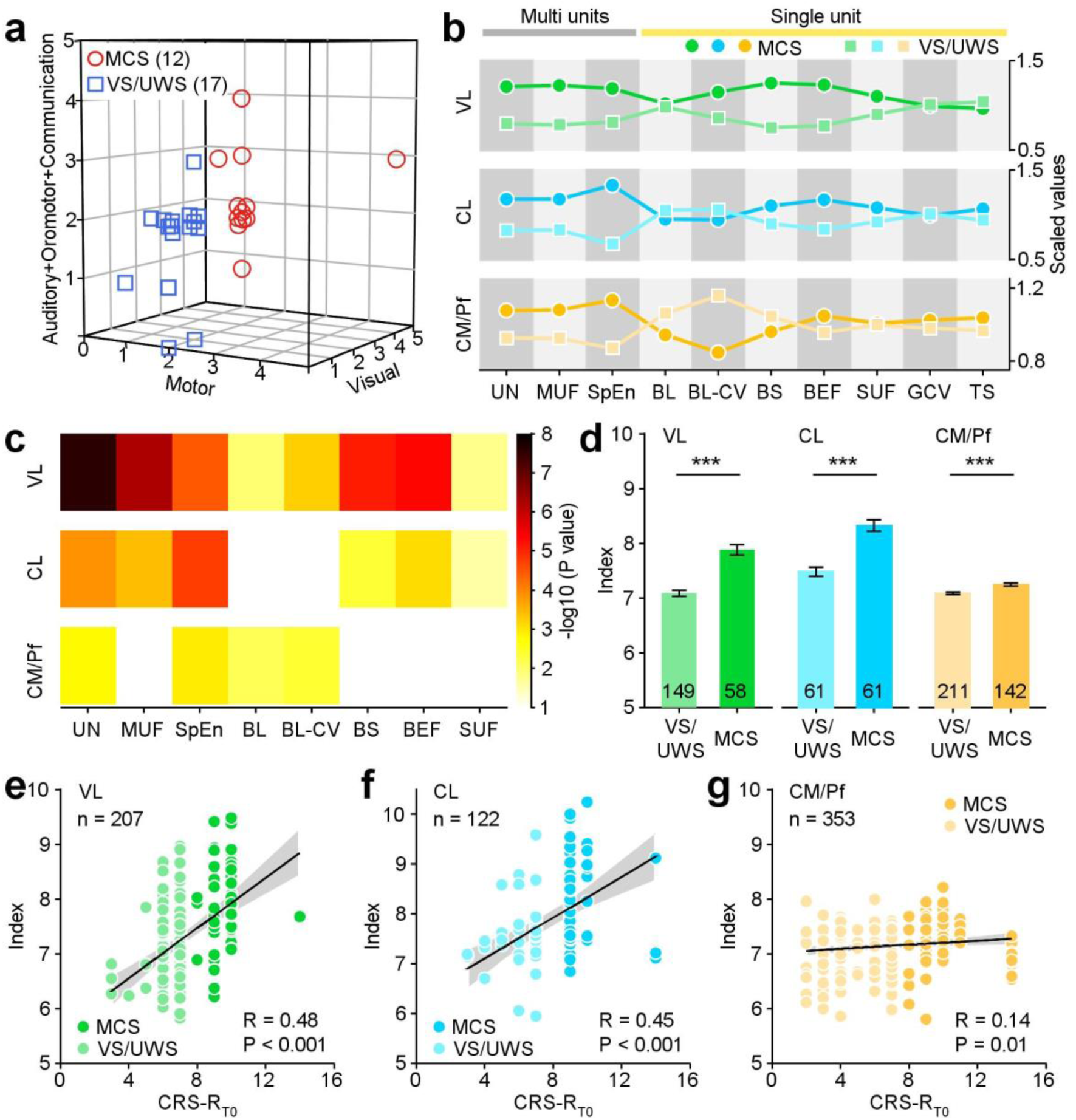
Neuronal signatures of thalamic neurons indicate DoC patients’ consciousness levels. **a**, 29 patients were diagnosed with MCS and VS/UWS regarding to the classic criteria^36^. **b**, The neuronal signatures in three thalamic nuclei are associated with DoC patients’ consciousness states. The scaled values of neuronal signatures were standardized by dividing the responses of each consciousness state by the mean responses across patients. **c**, MCS and VS/UWS have different neuronal signatures of the thalamic nuclei, according to Mann-Whitney-Wilcoxon tests. The colormap illustrates *P* values. **d**, Comparisons of neuronal indices of three thalamic nuclei between MCS and VS/UWS (***, *P* < 0.001, Mann-Whitney-Wilcoxon test, error bars: ±SEM). Number in each bar presents the number of recording sites. **e-g**, Positive correlations were observed between neuronal indices of individual recording sites in three thalamic nuclei and CRS-RT0 total scores (**e**, VL; **f**, CL; **g**, CM/Pf). Black lines represent the linear regression, while shaded areas are the 95% confidence interval of fitting.

To synthesize these signatures, we incorporated them as predictor variables in a partial least squares regression (PLS) model by applying the mean responses across neurons (see Methods for details). Based on the regression results (Extended Data Fig. 2a), we defined a neuronal index which can differentiate between patients’ consciousness states (Fig. 2d, *P* < 0.001, Mann-Whitney-Wilcoxon test). Further, the indices of three thalamic nuclei exhibit consistent trends of upward slopes as a function of patients’ total CRS-R scores (CRS-RT0, Figs. 2e-g). CRS-R scores were lower when indices were smaller and became progressively higher as indices grew larger. The neuronal index of the thalamus can differentiate MCS from VS/UWS and correlate with consciousness levels. Clinical factors, such as the interval injury to DBS surgery, as well as the etiology, could impact neuronal activities and are likely to play a major role in the manifestation of neuronal indices. It is noteworthy that the neuronal indices of thalamic nuclei consistently grade patients’ consciousness levels, employing a linear regression analysis to control for the effects of interval injury and etiology (Extended Data Figs. 2b,c).

### Thalamic spiking indicates arousal regulations following DBS in DoC patients

After a 7-day recovery from surgery, the thalamic DBS was delivered through the C0 contact of electrodes centered in the CM/Pf (Extended Data Fig. 1). As the direct treatment site, we examined whether neuronal signatures of the CM/Pf signal outcomes following DBS. We assessed patients’ recovery outcomes within 12 months using Glasgow Outcome Scales (GOS) and monitored their follow-up consciousness states by CRS-R scores (CRS-RT12). Ten patients recovered consciousness (CR, GOS ≥ 3). Their follow-up total CRS-R scores increased from 9 ±3 (Mean ± S.D., CRS-RT0) to 18 ± 4 (CRS-RT12). The remaining 19 patients did not recover consciousness (CNR, GOS ≤ 2); their total CRS-R scores evolved from 6 ± 2 (CRS-RT0) to 7 ± 2 (CRS-RT12). Neuronal activities of the CM/Pf in the CR patients revealed higher tonic firing rates and variations of ISI in single units, as well as more active neurons with higher spiking activities in multi-unit analyses (Figs. 3a-f, Mann-Whitney-Wilcoxon test, TS: *P* = 1.55 × 10^-4^; GCV: *P* = 8.63 × 10^-5^; SUF: *P* = 5.56 ×10^-3^; MUF: *P* = 9.18 ×10^-4^; SpEn: *P* = 1.53 ×10^-2^; UN: *P* = 6.47 ×10^-3^).

**Fig. 3.**
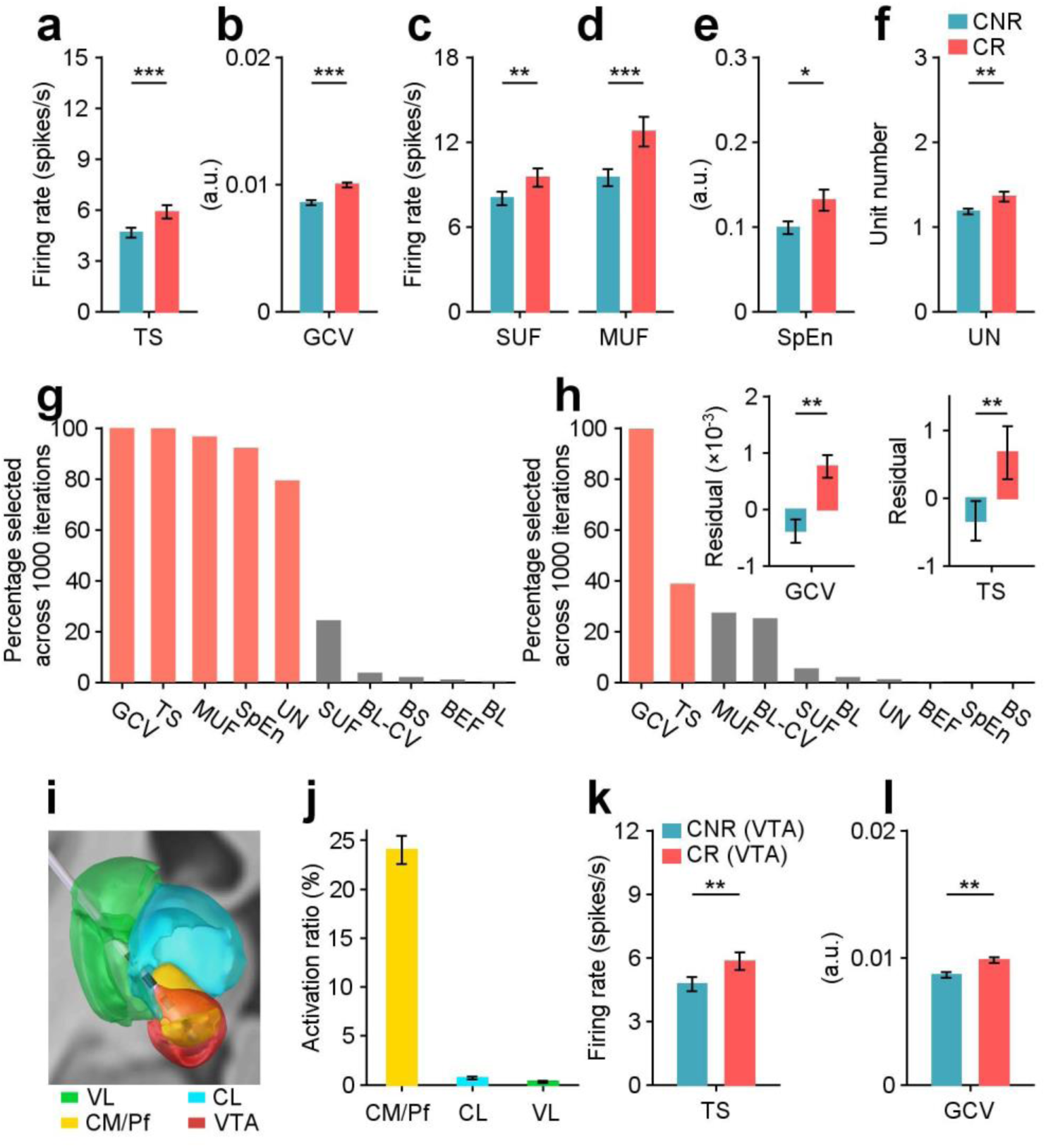
Thalamic neuronal signatures of the CM/Pf indicate arousal regulations following DBS in DoC patients. **a-f**, Comparisons of TS, GCV, SUF, MUF, SpEn and UN of the CM/Pf between CR (119 recording sites from 10 patients) and CNR (234 recording sites from 19 patients) groups. *, *P* < 0.05; **, *P* < 0.01; ***, *P* < 0.001, Mann-Whitney-Wilcoxon test. Error bars: ± SEM. **g**, Five top neuronal signatures of the CM/Pf were able to differentiate between CR and CNR (defined by F values, ANOVA), by using a cross-validated supervised machine learning model. **h**, Elimination the impact of patients’ preoperative consciousness states on neuronal signatures by a linear regression. GCV and TS of the CM/Pf consistently differ CR and CNR effectively. **i**, Estimation of the DBS VTA (red area) through the C0 contact in the example DoC patient (shown in **Fig. 1b**). **j**, Activation ratios represent the proportion of the VTA within each nucleus across patients, relative to the total volume of that nucleus. **k**, **l**, Comparisons of TS (**k**) and GCV (**l**) of recording sites located within the VTA area of the CM/Pf, between the CR and CNR groups.

We identify how these six neuronal signatures in the CM/Pf contribute to discriminate the recovery group from the non-recovery group by using a cross-validated supervised machine learning model (Fig. 3g, see Methods for details). The model indicates that five of them (except SUF) can sensitively signal recovery outcomes (CR vs. CNR) in 29 patients following DBS (defined by F-score, ANOVA). Prior to DBS, the average consciousness level in group CR (8 patients in MCS and 2 patients in VS/UWS) was notably higher than that of group CNR (4 patients in MCS and 15 patients in VS/UWS), potentially leading to varied recovery outcomes. Thus, these five thalamic signatures identified here might only serve as biomarkers to represent consciousness levels, rather than playing roles in arousal regulation or predicting outcomes. We ruled out this possibility by employing a linear regression analysis to control the effect of CR and CNR patients’ preoperative consciousness states. The residual results after regression revealed that the single-unit spiking activity could indicate recovery outcomes following DBS, including the single-unit tonic firing, and its ISI variation (Fig. 3h, Mann-Whitney-Wilcoxon test, TS: *P* = 1.42 ×10^-3^; GCV: *P* = 3.27 ×10^-3^).

In order to evaluate whether the neurons analyzed here were directly modulated by DBS, we simulated the DBS volume of tissue activated (VTA) in three thalamic nuclei and calculated their activation ratios by dividing the VTA by the total volume of that nucleus (Fig. 3i). Figure 3j depicts that 24.0% of the CM/Pf, 0.7% of the CL, and 0.3% of the VL could be activated by DBS in 29 patients. The existence of VTAs in the CL and the VL raised the possibility that DBS-activated neurons in these two nuclei could also aid in recovery following DBS. Comparing the recovery outcomes of patients with more than 1% or 2% activation ratios in the CL and the VL versus those without, our data failed to support this possibility. Neither GOS scores (1%: CL, *P* = 0.54, VL, *P* = 0.82; 2%: CL, *P* = 0.83) nor changes of total CRS-R scores following DBS (1%: CL, *P* = 0.37, VL, *P* = 1.00; 2%: CL, *P* = 0.74) permitted to differentiate both groups. Thus, we focused on the CM/Pf’s neurons located in the VTA regions of CR and CNR groups. The data showed that there was higher single-unit tonic firing (TS, Fig. 3k), as well as higher variation of ISI (GCV, Fig. 3l) in the CR group. Note that, as prospective prognostic indicators, TS and GCV of single units in the CM/Pf did not permit to differentiate consciousness levels (Fig. 2c). These findings suggest that neuronal mechanisms for altering the levels of consciousness may be distinct from neuronal signatures involved in representing the states of consciousness.

### Thalamic neuronal signatures linked with thalamocortical connections to predict recovery outcomes

The thalamus assumes the responsibility of regulating arousal through its connections within the neuronal network, which include the cerebral cortex and striatum. We examined a subgroup of 17 patients who obtained qualified rs-fMRI data and conducted three independent additional analyses to reveal the relationships between thalamic signatures, brain connections and DBS outcomes (Fig. 4a).

**Fig. 4.**
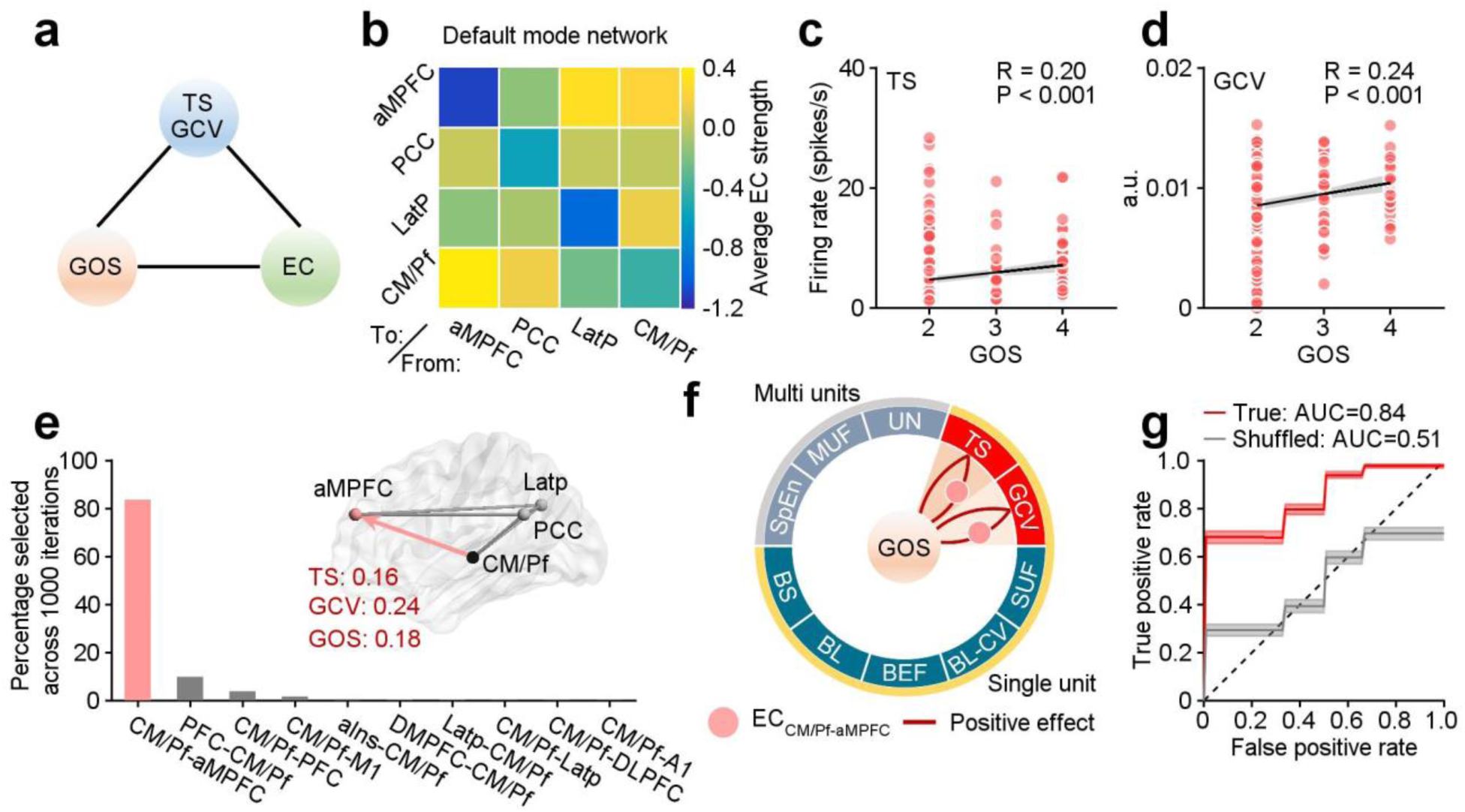
DoC patients’ thalamic firing linked with thalamocortical connections to predict outcomes following DBS. **a**, Schematic diagram of pairwise relationships between neuronal signatures, GOS scores, and ECs. **b**, Effective connectivity between the CM/Pf and cortical areas in the default mode network, averaging across 17 patients with qualified rs-fMRI data. **c**, **d**, Correlation between neuronal signatures of 306 recording sites from 17 patients (TS: **c**, and GCV: **d**) and GOS scores, measured by Pearson’s correlation coefficient. The solid lines represent linear regression, whereas shaded areas represent 95% confidence intervals. **e**, The top connection selected by F values of ANOVA test was able to discriminate the CR group from the CNR group in our dataset. The thalamocortical connection from the CM/Pf to the aMPFC was positively correlated with GOS scores and thalamic neuronal signatures, with related effect sizes listing in red fonts. **f**, Summary of relationships between thalamic signatures, ECs and DBS outcomes in DoC patients. The ring contains ten neuronal signatures of the CM/Pf. Seven of them were defined from single units (yellow arcs), whilst the remaining three signatures were estimated from multiple units (gray arcs) of each recording site. Pink spots present the ECCM/Pf-aMPFC. Lines indicate “logic” loops of correlations between neuronal signatures (TS and GCV), the ECCM/Pf-aMPFC, and GOS scores, with red lines representing positive effects. **g**, Receiver operating characteristic curves to predict recovery outcomes based on the top neuronal signatures (TS and GCV) and the top connection (ECCM/Pf-aMPFC) from individual patients, reflecting the Mean ±SEM over 1000 times for the true (red) and shuffled (gray) models.

Initially, the relationship between effective connectivity (EC) and recovery outcomes were examined. We utilized Dynamic Casual Modeling (Fig. 4b and Extended Data Figs. 3a-f) to investigate the thirty connections between the CM/Pf and 15 cortical areas from 6 functional brain networks, namely the default mode network, the executive control network, the salience network, the sensorimotor network, the auditory network, and the visual network, as well as two connections between the CM/Pf and the striatum (Extended Data Fig. 3g). The subsequent examination aimed to investigate the correlation between neuronal signatures in the CM/Pf and recovery outcomes. Upon reevaluating the neuronal signatures in these 17 patients with qualified rs-fMRI data, the findings consistently demonstrated that both TS and GCV in the CM/Pf were correlated with GOS scores (Fig. 4c,d). Lastly, we investigated how neuronal signatures of the CM/Pf influence brain connections, by utilizing the Parametric Empirical Bayes analyses on neuronal signatures and effective connectivity (Fig. 4e).

The three pairwise correlation analyses discovered 6 ECs that formed “logic loops”. These loops establish logic linkages between ECs, neuronal signatures, and GOS scores in a closed manner (Extended Data Figs. 4a,b). Intriguingly, these ECs comprised the anterior medial prefrontal cortex (aMPFC), superior parietal cortex (SPC), dorsal anterior cingulate cortex (dACC), primary auditory cortex (A1) and anterior prefrontal cortex (PFC). Most are crucial nodes in consciousness and wakefulness-related thalamoparietal and thalamofrontal connections. In contrast, the connections between the CM/Pf and the striatum failed to yield any statistically significant differences between the CR and CNR groups (Mann-Whitney-Wilcoxon test; ECCM/Pf-Striatum: *P* = 0.19; ECStriatum-CM/Pf: *P* = 0.28). The extensive thalamostriatal connections in both CR and CNR patients might have potentially prevented accurate predictions of recovery.

To further verify the impact of these brain connections on recovery outcomes, we utilized the cross-validated supervised machine learning model with a feature selection. The analysis has evaluated the most important connection that effectively differentiates the CR group and the CNR group (Fig. 4e): the EC from CM/Pf to aMPFC in the default mode network (ECCM/Pf-aMPFC). Within the logic loops in Figure 4f, higher neuronal tonic firing rates and variations of ISI in the CM/Pf are correlated with better recovery outcomes with higher GOS scores and the strengthened ECCM/Pf-aMPFC. To summarize these effects, we utilized a logistic regression classification to predict DBS outcomes. Results from the model demonstrate that the top selected factors, including TS, GCV, and ECCM/Pf-aMPFC of individual patient, effectively discriminate the CR group from the CNR group, achieving an AUC of 0.84 (Fig. 4g). These relationships embedded in a logic linkage indicate that neuronal signatures (TS and GCV) in the CM/Pf could impact recovery by modulating the effective connectivity (ECCM/Pf-aMPFC). It suggests that the CM/Pf, as a location for DBS, may be an origin of effects on regulating arousal in neuronal networks.

## Discussion

In this study, we have been able to evaluate cellular events that occur in the thalamic nuclei of patients diagnosed with VS/UWS and MCS, at both single neuron and multiple neuron levels. Our results demonstrated effects of thalamic spiking activity on consciousness levels in patients with DoC. In patients with a higher consciousness level, larger spiking activities, more active neurons, and more asynchronous firing patterns (higher SpEn) in the thalamus could generate stronger neuronal outputs and more flexible connections. These effects were consistent with previous research on neuronal activities in the healthy brain when animals woke from anesthesia and sleep^7,10,37^. In addition, our data revealed that the burst discharge of single neurons was an essential signature for the representation of consciousness in DoC patients. Patients in a VS/UWS who experience a lower level of consciousness demonstrated longer and more variable burst discharges in the CM/Pf compared to those with a higher consciousness in a MCS. Previous research demonstrated that shorter burst lengths are indicative of a stronger γ-aminobutyric acid-ergic (GABAergic) inhibition in the system, as the burst length can be shortened by injecting GABA into striate cortex and prolonged by applying GABAA receptor blocker bicuculline^38^. DoC patients with traumatic brain injury showed broad reductions in GABAA receptor availability predominantly in the frontal lobes, striatum, and thalamus^39^, which could result in longer burst lengths. Neuronal burst discharges in the cortex and subcortex may share common mechanisms for consciousness-related processing. By combining these effects, our PLS regressions on thalamic signatures defined an index for assessing consciousness levels in DoC patients. Intriguingly, neuronal indices of thalamic nuclei can sensitively grade distinct levels of consciousness, ranging from VS/UWS to MCS.

Recent neuroimaging studies have unveiled intricate functional alterations within widespread brain networks of DoC patients. One of the prominent findings is the disrupted default mode network (DMN), which comprises brain regions implicated in introspection, self-referential processing, and mind wandering^40–47^. Consistent reports indicate reduced connectivity within the DMN and altered interactions with other brain networks in DoC, suggesting a breakdown in intrinsic brain activity and consciousness mechanisms^4,48–55^. As a key node of the DMN, the aMPFC had specific role in cognitive functions^56^. Research indicates that both the connection features of the aMPFC and its functional connectivity with the dorsomedial prefrontal cortex correlate with patients’ total CRS-R scores, offering potential as predictors of recovery outcomes^57^. Studies further reveal variations in functional connections involving the aMPFC across different sleep states^58^ and attenuation under sedation^59^, shedding light on its role in consciousness. Additionally, impaired thalamocortical connection emerges as another relevant finding in DoC patients. The anterior forebrain mesocircuit underscored the essential roles of thalamus in consciousness alterations^1^, through extensive connections with the frontal and parietal cortex (^8,60–65^, our data in Extended Data Figs. 3h,i). Our results revealed that the connections between the CM/Pf of the thalamus and the cortex correlate with recovery outcomes following DBS (Extended Data Fig. 4). Previous research on anesthetized animals showed that DBS in the CM/Pf can effectively affect cortical networks. CM-DBS^11^ and CM/Pf-DBS^32^ modulated the activity in various brain regions, including the prefrontal, parietal, cingulate, temporal, and occipital cortices, as well as the striatum. Furthermore, DBS restored the collapsed principle gradient of functional connectivity and the reduced network hierarchical integration induced by anaesthesia^33^. This resulted in arousal-related functional responses in non-human primates. Significant sensorimotor-associative and limbic-associative BOLD effects were produced by CM- and Pf-DBS, respectively^66^. Our study demonstrated that there were bidirectional brain connections inside logic loops linking to recovery outcomes following DBS: ECs from the CM/Pf to the cortex (Extended Data Fig. 4a), and ECs from the cortex to the CM/Pf (Extended Data Fig. 4b). When comparing the neuronal signatures of TS and GCV, we observed that tonic firing only impacted the unidirectional connection from the CM/Pf to the cortex, rather than the opposite direction (Extended Data Fig. 4c). These findings hold essential implications for understanding the organization of neuronal mechanisms underlying thalamic DBS in regulating arousal. As a direct DBS site, neuronal signatures of TS and GCV in the CM/Pf, as well as ECs between the CM/Pf and the cortex exhibit correlations with GOS scores, suggesting their potential in predicting recovery outcomes. Therefore, based on the effect on unidirectional connections, higher tonic firing in the thalamus may be amenable to DBS modulation through modulating the connection from the CM/Pf to the cortex, especially ECCM/Pf-aMPFC (Fig. 4).

It is the context of thalamic firing we demonstrate that is most noteworthy. Signatures of single units in the CM/Pf associated with recovery outcomes are distinct from those grading consciousness levels. We have thoroughly analyzed dual modes of thalamic neurons: burst and tonic discharges. Burst events are crucial for discriminating VS/UWS patients from MCS patients, whereas tonic discharges transmit effective signals for altering consciousness levels following DBS. Rapid switching of thalamic firing modes (burst or tonic) in mice can govern the transition of sleep-wake states^9^ or the generalization of epilepsy^67^ in a bidirectional manner: burst firing triggered slow-wave-like activity or initiated absence epilepsy, whereas tonic activation promoted sleep recovery or terminated the symptom. Thalamus could be the key center for the representation and alteration of consciousness, but via distinct neuronal mechanisms. The burst and tonic discharge of thalamic neurons appear to play crucial roles for each of them.

A growing body of research indicates that the CL could play a crucial role in arousal regulation^8,10,68^. CL-DBS could alter consciousness by enhancing interactions in the frontal eye field and the lateral intraparietal area^10^. We have evaluated the CL’s potential contribution to DBS effects in these patients. Five neuronal signatures of the CL showed differences between the CR and CNR groups (Extended Data Figs. 5a-e, Mann-Whitney-Wilcoxon test, *P* < 0.05). By utilizing a linear regression analysis to control patients’ preoperative consciousness states, our data verified that only the GCV in the CL could be consistently selected to discriminate the CR patients from the CNR group in our dataset (Extended Data Fig. 5f). The thalamocortical connections of ECCL-alns and ECCL-aMPFC were identified as the most effective features to discriminate the CR and CNR groups (Extended Data Fig. 5g). Nevertheless, the GCV of CL failed to be significantly related to neither brain connections nor GOS scores (Extended Data Fig. 5h), which cannot support the logic loops (Extended Data Figs. 5i,j). Possible reasons could include the presence of a limited number of recording sites in the CL along the microelectrode punctures and a scarcity of electrically activated volumes in the CL (Fig. 3g) by DBS in these patients.

The question of how the thalamus modulates consciousness is answered in the most detail for the representation of consciousness and effects on arousal regulation using DBS in a group of 29 patients with DoC. We anticipate that these details may pave the way for the development of targeted interventions and prognostication strategies for affected individuals. As a potential predictor of DBS outcomes, stronger thalamic tonic firing could be considered as one of important factors to be evoked when devising stimulation parameters for therapeutic interventions. In the meantime, it is almost certain that the thalamic principles of consciousness will apply to other brain diseases. Perhaps epilepsy, Parkinson’s disease, and their DBS outcomes could also be signaled by thalamic spiking activity and thalamocortical connections.

## Methods

### DoC patients

This retrospective research included a total of 29 patients who were confirmed with DoC and received thalamic DBS as their clinical treatments (Extended Data Fig. 1). They were diagnosed in a MCS (12 patients) or a VS/UWS (17 patients) by neurosurgeons according to CRS-R scores.^36^ These patients exhibited prolonged DoC for a minimum of 28 days after brain injury, without serious structural brain damage or abnormalities. The preoperative consciousness states were scored as CRS-RT0 (Extended Data Fig. 1a). Follow-up assessments were normally scheduled at four intervals after the DBS surgery: 1 month, 3 months, 6 months and 12 months. The GOS^69^ was used as the main prognostic assessment, and the follow-up CRS-R was assessed to monitor consciousness states following DBS.^57^ Patients who showed ongoing improvement were evaluated based on the 12-month follow-up outcomes. Data from the most favorable follow-up assessment were considered for patients whose condition deteriorated due to complications. Both GOS and CRS-R scores of patients were assessed by experienced neurosurgeons among the 12-month follow-up assessments (T12 in Extended Data Fig. 1a). Extended Data Table 1 detailed patients’ information.

The clinical dataset analyzed in the study was acquired from DoC patients undergoing clinical treatments at the Neurosurgery Departments of two hospitals. The study was approved by the Ethics Committee of Beijing Tiantan Hospital, Capital Medical University (protocol No: KY2017-361-01) and the Ethics Committee of PLA Army General Hospital (protocol No: 2011-0415). The patient’s parents or legal guardians signed two patient consent forms after receiving thorough information and discussion about the treatment processes. One consent form provided comprehensive details about the treatment processes, outlining the surgical procedure, accompanying risks, and the potential for the treatment to be entirely ineffective. The other consent form pertained to research participation, stating that patients’ data obtained before, during and after surgery (such as MRI, CT, intraoperative thalamic spiking recordings and behavioral assessments) would be utilized for prospective research analyses. Both consent forms ensured that patient’s parents or legal guardians were fully informed and gave consent to aspects of the treatment and potential prospective research. All clinical data utilized in this study were examined and gathered to serve for clinical treatments. There were no extra examinations attached for research purposes.

### Blinding

The collection and analyze of patients’ data were conducted by separate groups of investigators in a completely blinded fashion. Investigators from two Neurosurgery Departments who collected clinical data were blinded to the properties of functional brain connectivity in DoC patients and their full sets of thalamic spiking signatures. Investigators from the Institute of Biophysics who performed the analyses on thalamic spiking profiles and the brain connections were blinded to patients’ states of consciousness and the outcomes of DBS until they finished all the spiking signatures and brain connections analyses.

### MRI acquisition

Prior to surgery (T0, Extended Data Fig. 1a), patients received resting-state functional magnetic resonance imaging (rs-fMRI), T1-weighted 3D high-resolution and diffusion MRI scanning. During the MRI scan, no sedatives or anesthetics were administered to the patients. Rs-fMRI scans were acquired using a T2-weighted gradient echo sequence and diffusion MRI scans were acquired using a EP/SE sequence on 3.0 T scanners. Details of scanning information were listed in Extended Data Table 2.

### DBS implantation and reconstruction of lead trajectories during surgery

The surgical planning was designed by *Leksell SurgiPlan* for utilization with the *Leksell* stereotactic system (Elekta, Stockholm, Sweden). The anatomical location of the CM/Pf was 7.8-9.7 mm posterior to the middle of the anterior commissural (AC)-posterior commissural (PC) line, 8.8-10.5 mm distal to the AC-PC line (4.5-5.5 mm from the ventricular wall), and 0-1.5 mm inferior to the AC-PC plane^28^. Twenty-seven patients had bilateral implantation of quadripolar DBS leads (Medtronic #3387, USA, or PINS L302, China), while two patients had unilateral implantation, for a grand total of 56 trajectories (Extended Data Fig. 1c). Microelectrode recordings were employed during surgery when patients recovered from anesthesia to help determine the targeting location of the CM/Pf, as this required the use of considerably less distracting background noise and neuronal activities^28,70^. The thalamic activities, as observed through single-unit and multi-unit frequencies in our data, exhibited a decrease when microelectrodes were introduced through VL, CL to CM/Pf (Extended Data Figs. 1d,e). Then, DBS leads were implanted into the thalamus along the same path as the microelectrodes. Standard punctures involved microelectrodes and leads entering the thalamus at the VL, traversing the CL, and aiming for a region between the CM and PF (Fig. 1b).

To verify the accuracy of their placements, postoperative CT or MRI images were utilized to reconstruct lead trajectories applying Lead-DBS software version 2.5.3 (https://www.lead-dbs.org/)^71^. Using Advanced Normalization Tools (ANTs)^72^ and Statistical Parametric Mapping (SPM12)^73^, respectively, postoperative CT and MRI brain images were co-registered to preoperative MRI. Coarse subcortical mask^74^ was conducted (as implemented in Lead-DBS) to eliminate bias generated by brain shift that might occurred during surgery. Images were nonlinearly normalized to the Montreal Neurological Institute (MNI, ICBM 2009b nonlinear asymmetric) template space using the SyN Diffeomorphic Mapping approach implemented in ANTs, with the preset “Effective: Low Variance + subcortical refinement” to acquire a most precise subcortical alignment between patients and MNI space. This method was considered to have the best performance on subcortical registrations, which was proposed by a recent comparative study^75^ evaluated 6 different nonlinear atlas-based subcortical normalization and segmentation methods, and reproduced independently by another study^76^. The trajectories were then reconstructed automatically using the TRAC/CORE approach and manually modified to evaluate the contact points with more precision. Hence, lead trajectories were reconstructed and contact coordinates were transformed into MNI space.

### DBS stimulation and the volume of tissue activated (VTA) modeling

The DBS stimulation was administered seven days after the operation, after the incision had healed and the edema generated by the puncture had subsided. Periodic electrical stimulation was delivered via C0 contact to the CM/Pf of patients (Extended Data Fig. 1c). The primary stimulation source was monopolar and consisted of 100 Hz, 120 μs, and 3.0-4.0 V^28,77^. Continuous stimulation was administered from 8 AM to 8 PM with a cycle of 15 min on and 15 min off (Extended Data Fig. 1b).

To evaluate effective activation fields of DBS, we simulated the volume of tissue activated in patients’ native space based on contact configuration and stimulation voltage settings by using a finite element method^71^. A volume conductor model was conducted based on a fosur-compartment mesh that separated perielectrode tissue into gray matter (nuclei were defined by Krauth/Morel thalamic atlas^78^), white matter, electrode conducts and insulated parts. Then, the activation field distribution was simulated using an adaptation of the FieldTrip-SimBio pipeline implemented in Lead-DBS and transformed to MNI space using the aforementioned normalization warp fields. To define the binary activation field, a threshold of 0.2 V/mm^71^ was applied. We simulated activation fields with voltages ranging from 3.0 to 4.0 V and obtained similar results. The data with a voltage setting of 3.5 V were demonstrated in Figure 3f.

### Intraoperative microelectrode recordings (MERs)

Anesthesia with the propofol was terminated 30 minutes prior to MERs during surgery. Microelectrodes with a single channel were used intraoperatively to record neuronal activities (LeadPoint System, Medtronic, USA; Neuro Nav, Alpha Omega, Israel). Signals were sampled at 24 kHz and subjected to hardware-based bandpass filtering (500-5000 Hz) and notch filtering (50 Hz). Each site was recorded for 10 seconds. MER acquisitions generally began 10-11 mm above the target and advanced in 0.5 mm steps to a depth of 1-3 mm below the target.

The anatomical position of recording sites was identified based on their depths in millimeters (relative to the target position) along the lead trajectories reconstructed. This resulted in three-dimensional coordinates for MERs in both brain hemispheres. According to the lowest Euclidean distance between recording sites and masks from the Krauth/Morel thalamic atlas, recording sites were allocated to the thalamic nuclei from whence they originated^78^. 731 recording sites located in the thalamus from 29 individuals were included in the subsequent spike sorting analysis.

### Spike sorting

We used a Wave_clus MATLAB tool (https://github.com/csn-le/wave_clus)^79^ for spike identification and sorting with default parameters. The processes of this algorithm are outlined below. The raw MER data were zero-phase filtered with a fourth-order bandpass elliptic filter between 500 and 5000 Hz. Spikes were detected with an automatic amplitude threshold, which was set as 5 times the estimated standard deviation of the noise. During the feature extraction phase, wavelet coefficients of each detected spike were retrieved using a four-scale multiresolution decomposition with a Haar wavelet. Ten wavelet coefficients with the greatest capacity for distinguishing different spike waveform shapes were chosen through a Lilliefors modification of the Kolmogorov-Smirnov Test of Normality. At last, an unsupervised super-paramagnetic clustering approach was used performed to classify spike waveform characteristics into different clusters based on the 11-nearest neighbor interactions. The main parameter that affect the performance of super-paramagnetic clustering is the ‘temperature’. This parameter’s range was set from 0 to 0.25 in steps of 0.01. Optimal temperature was determined as the highest temperature when a cluster contained at least 20 members. We calculated the signal-to-noise ratio (SNR) of single unit as the amplitude of mean waveform (trough-to-peak) divided by the standard deviation. ISI violation percentage was also calculated as the percentage of all spikes with ISI smaller than 1 ms. Only units with SNR greater than or equal to 2 and ISI violation less than or equal to 1 % were used in further analyses. Spikes within 1 ms ISI were removed from spike trains.

Using the spike sorting approach, we have gathered 1016 neurons from a total of 731 thalamic recording sites. 94.49% of them (960 out of 1016) exhibit two distinct firing modes, burst and tonic. The subsequent investigations of neuronal signatures focused on these 960 neurons from 682 sites.

### Single unit and multi units

We evaluated neuronal signatures of the 682 recording sites based on their single unit and multi-unit activities: 207 sites were in the VL (including anterior and posterior subdivisions), 122 sites were in the CL, and 353 sites were in the CM/Pf (Fig. 1c). There were multiple neurons recorded at 42.51% (88/207), 31.97% (39/122) and 18.41% (65/353) of the recording sites in these three thalamic nuclei. The single-unit studies included analyses of single-neuron firing rate, burst, and tonic activity. In order to mitigate potential bias towards specific types of thalamic neurons at each recording site, we conducted analyses on the mean responses across neurons to present signatures on the single-neuron level if there were multiple units recorded at a given site. The multi-unit firing rates, neuron numbers, and sample entropy were utilized to describe the neuronal signatures of multiple units at that site. The neuronal firing rates were computed by dividing the amount of spikes by the recording duration.

### Burst and tonic modes

To capture the temporal properties of neuronal firing, we generated the inter-spike interval histograms with a bin width of 0.5 ms for each single unit using FieldTrip software (https://www.fieldtriptoolbox.org/)^80^. The burst mode was defined by clusters of at least two spikes with ISIs less than 8 milliseconds (Fig. 1f). We calculated burst frequency (BF, bursts per second), burst length (BL, spikes per burst) ^38^, and burst length’s coefficient of variation (BL-CV). In addition, we established burst events by considering with ISIs that were less than 5 ms and 10 ms, and obtained comparable results.

### Sample entropy (SpEn)

Sample entropy was applied to assess the regularity and complexity of time series. It is theoretically similar to approximate entropy, but more precise^81^. A low value of SpEn often indicates a high degree of regularity^37,82^. Here, we utilized SpEn to estimate the irregularity of multi-unit activities. Spike trains of multi units were convolved with a Gaussian kernel having a standard deviation of 25 ms (FieldTrip function ft_spikedensity) in order to obtain continuous signals, and then down sampled to 125 Hz^37^. To calculate the SpEn, a time series of finite length *N*, *x*(*i*), 1 ≤ *i* ≤ *N*, was first embedded with a dimension of *m*:

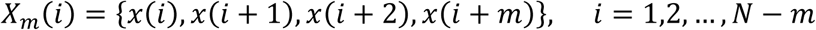

Then, the probability that *X_m_*(*j*) is within distance *r* of *X_m_*(*i*) was defined as follows:

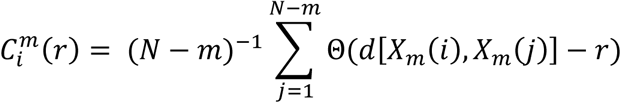

where Θ is the Heaviside function and *d* is the Chebyshev distance between *X_m_*(*i*) and *X_m_*(*j*). The embedding dimension *m* was set as 3 and distance criteria *r* was set as 0.2 SD of continuous signals of multi units^37^.

Finally, the SpEn was defined as:

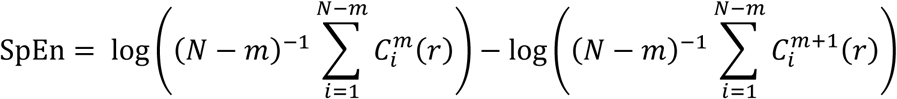

The measure was implemented by the MATLAB function SpEn (files owned by Kijoon Lee; https://ww2.mathworks.cn/matlabcentral/fileexchange/35784-sample-entropy).

### Geometric coefficient of variation (GCV)

In order to evaluates the variability in spiking time of each single neuron, we used the following formula to quantify the GCV^83^ of ISIs below 40 ms:

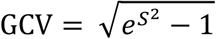

where *S* is the standard deviation of ISIs. GCV is high when spikes are more sparsely distributed, it is low when spikes are clustered as burst event with regular ISIs.

### Partial Least Squares (PLS) regression analyze and neuronal index definition

PLS regressions were performed to predict consciousness levels based on patients’ CRS-RT0 and neuronal signatures (independent variables). Only those signatures (four signatures in the CM/Pf, six signatures in the CL, and eight signatures in the VL) passed the statistical test between MCS and VS/UWS groups were applied in the PLS regression analyses (Fig. 2c, Matlab function plsregress). The PLS model returned the regression coefficient of each neuronal signature and the intercept term (Extended Data Fig. 2a). Then, the neuronal index was defined using the following formula:

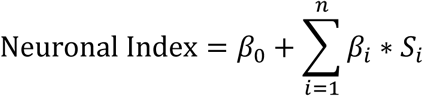

where β_0_ is the intercept term, β_i_ is the regression coefficient of the ith neuronal signature S, n=8 in the VL, n=6 in the CL, and n=4 in the CM/Pf.

### Resting-state image preprocessing and effective connectivity analyses

In this study, 18 of 29 patients were accessible for rs-fMRI Analysis. fMRI preprocessing and connectivity analyses were carried out as described in previous research^57^. For the analysis of resting-state fMRI images, SPM12 (https://www.fil.ion.ucl.ac.uk/spm/software/spm12/) and freely accessible programs (https://github.com/elifesciences-publications/pDOC) were utilized. The preprocessing steps comprised the elimination of the initial five volumes, slice timing, head motion correction, spatial smoothing with a 6-mm Gaussian kernel, nuisance signal regression, and temporal bandpass filtering (0.01-0.08 Hz). During the nuisance signal regression, linear regression was utilized to eliminate the impact of head motion (12 motion parameters, including roll, pitch, yaw, translation in three dimensions and their first derivatives), whole brain signals and linear trends. To minimize the effects of motion artifact on functional connectivity analysis, framewise displacement of head movement was evaluated, and volumes with large movements also were removed. It is the sum of the absolute values of the translational and rotational realignment estimates’ derivatives (after converting the rotational estimates to displacement at 50 mm radius)^84^. Volumes with framewise displacement more than 1.5 mm were discarded, and patients with less than 50 remaining volumes were excluded for further analysis. According to these criteria, one (No. 08) of 18 patients with rs-fMRI was discarded. The subsequent effective connectivity analyses were conducted using the 17 patients’ rs-fMRI data.

Next, we utilized the dynamic casual modeling (DCM) module of SPM12 to examine the effective connectivity between thalamic nuclei (CM/Pf and CL) and cortical regions in six brain networks (the default mode, executive control, salience, sensorimotor, auditory, and visual networks, Extended Data Figs. 3a-f), as well as between thalamic nuclei and the striatum (Extended Data Fig. 3g). The default mode network includes the anterior medial prefrontal cortex (aMPFC), posterior cingulate cortex/precuneus (PCC), and lateral parietal cortex (LatP). The execute control network consists of the dorsal medial prefrontal cortex (DMPFC), the anterior prefrontal cortex (PFC), the superior parietal cortex (SPC), and the dorsal lateral prefrontal cortex (DLPFC). The orbital frontoinsula (Alns) and the dorsal anterior cingulate cortex (dACC) are salience network components. Sensorimotor network is composed of primary motor cortex (M1) and supplementary motor area (SMA). The primary auditory cortex (A1) and the middle cingulate cortex (MCC) constitute the auditory network. The primary visual cortex (V1) and the associative visual cortex (V4) make up the vision network. The ROIs of CM/Pf and CL were established by the Krauth/Morel atlas^78^. The ROIs of cortical regions within six brain networks were identified by the methods developed by Song et.al^57^. The ROIs of striatum were established by the atlas of the basal ganglia (ATAG)^85^. The combination of voxels from both hemispheres produced the thalamic ROI and fourteen cortical ROIs. The first principal component of the time series from each ROI was computed by SPM12 and saved as the volume of interest (VOI) for that ROI. Extracted VOIs were used in subsequent DCM analysis.

Each thalamocortical DCM model featured CM/Pf or CL and brain regions in a particular network. The effective connectivity includes the reciprocal connections between these brain regions, and their self-connections within each region. Due to the efficacy of the spectral DCM (sDCM) approach in analyzing rs-fMRI data^86^, these models were estimated for each patient using the sDCM method^87^. The estimated model parameters were averaged across patients using Bayesian fixed effect averaging method (spm_dcm_average). The group-level analyses were performed using the Parametric Empirical Bayes (PEB) method (spm_dcm_peb and spm_dcm_peb_bmc)^88^. The effects of neuronal signatures and GOS were evaluated by specifying them as the covariates of the second-level design matrix. The design matrix consisted of two columns of covariates: the first column was ones (to model the commonality effect across patients, i.e., the constant or group mean), and the second column represented the between-subject influences of neuronal signatures or GOS (mean-centered). Before being specified as a covariate, a given neuronal signature was averaged across CM/Pf neurons for each patient, and then the mean values of that signature were z-scored across patients. To focus on the parameters with stronger evidence, we applied a threshold to the Bayesian Model Average in PEB, selecting only those having a 99% posterior probability of being present vs. absent (thresholding based on the free energy, Figs. 4e,f; Extended Data Figs. 4a,b and 5i,j).

### Structure connectome

We analyzed structure connectome between the thalamic nuclei and cortical areas, as well as the striatum (Extended Data Figs. 3h,i), using diffusion MRI data of DoC patients, conducted by DSI-Studio (<u>DSI-Studio: A Tractography Software Tool for Diffusion MRI Analysis | DSI Studio</u> <u>Documentation</u> (labsolver.org)). After preprocessing (motion correction and removing eddy current distortion), the 1.25 mm isotropic resolution spin distribution function (SDF) of each patient was reconstructed to MNI space. This reconstruction was achieved using the Q-space diffeomorphic reconstruction (QSDR) method^89^, with a diffusion sampling length ratio of 1.25.

A deterministic fiber tracking algorithm was utilized to achieve whole brain tracking, tracking parameters were set as: angular threshold = 0, step size = 0, minimum length = 30 mm, maximum length = 300 mm, number of tracts = 1000000. The ROIs of cortical areas were defined using the HCP-MMP atlas^90^ built-in DSI-Studio, the ROIs of striatum were defined using the atlas of the basal ganglia (ATAG)^85^, and the ROIs of thalamic nuclei were defined using the Krauth/Morel atlas^78^. The connectivity matrix was calculated by considering the number of tracts that pass through paired ROIs. For each DoC patient and HCP subject, a threshold of 0.001^91^ (the ratio to the maximum connecting tracks in the connectivity matrix) was used to filter out the connectivity matrix. Connectivity matrices of individuals were averaged to obtain the average matrix which was also filter out using the threshold of 0.001.

### Feature selection and logistic regression analyses

Feature selection was conducted using the algorithm of f_classif implanted in Python library of Scikit-learn^92^, it computes the ANOVA F-value to select the most distinguishing features between consciousness recovery and non-recovery groups. Two independent classification analyses were separately performed on 10 neuronal signatures and 32 brain connections (30 connections between thalamic nuclei and cortical regions, 2 connections between thalamic nuclei and the striatum). Binary classifications were carried out with penalized logistic regression through the algorithm of LogisticRegression of Scikit-learn^92^. Briefly, the label *y_i_* of data point *i* takes values in the set [0, 1] (0 for consciousness non-recovery and 1 for recovery), and the probability of positive class was predicted as:

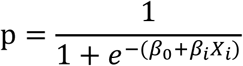

where β_0_ is the intercept of the logistic regression model, β*_i_* is the slop coefficients of the model, *X_i_* are features. To optimize model parameters, the L2 regularization term *r*(β) was combined to minimize the cost function:

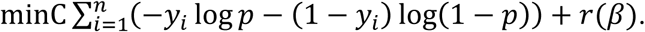

The logistic regression classification was cross-validated using the k-folds method (5 folds for neuronal signatures and 4 folds for effective connectivity) and repeated 1000 times to establish stability in feature importance histograms (Figs. 3g,h and Fig. 4e) and AUC scores (Fig. 4g). The shuffling analysis was conducted by randomly permuting class labels 1,000 times, and the output of the logistic classifier was compared with results of the shuffled data.

### Statistical analysis

The neuronal signatures and neuronal indices of MCS and VS/UWS patients (Figs. 2c,d), as well as neuronal signatures between the recovery and non-recovery groups (Figs. 3a-f, h, k, l and Extended Data Figs. 5a-e) were compared utilizing a Mann-Whitney-Wilcoxon test. The frequency of single-unit and multi-unit activity in the CM/Pf, the CL and the VL were compared using a one-way analysis of variance (ANOVA) test, followed by a post-hoc Tukey-Kramer test for pairwise comparisons between thalamic nuclei (Extended Data Figs. 1d,e). Statistical analysis was conducted to determine the comparisons, using a significance level of 0.05. Pearson’s correlation coefficient was used to measure the correlation between neuronal index and CRS-RT0 and between neuronal signatures and GOS scores (Figs. 2e-g; 4c, d; Extended Data Figs. 2b, c; 5h).

## Data availability

Datasets supporting the findings of this study are available from the corresponding authors on reasonable request.

## Code availability

A spike sorting toolbox Wave_clus was used to identify neuronal spikes (https://github.com/csn-le/wave_clus). The SPM12 (https://www.fil.ion.ucl.ac.uk/spm/software/spm12/) and freely accessible programs (https://github.com/elifesciences-publications/pDOC) were used to analyze rs-fMRI data. Trajectories of DBS leads were reconstructed using Lead-DBS software (https://www.lead-dbs.org/). The analysis of ISI and spike train convolution of multi units were performed by Fieldtrip software (https://www.fieldtriptoolbox.org/). PLS regression was achieved using the MATLAB function plsregress. The computation of SpEn was based on the MATLAB function SpEn (https://ww2.mathworks.cn/matlabcentral/fileexchange/35784-sample-entropy, files owned by Kijoon Lee). Feature selection and cross-validated logistic regression were conducted using Scikit-learn (https://scikit-learn.org). Custom codes used in the study are available from the corresponding authors on request.

## Acknowledgments

This study was supported by Beijing Natural Science Foundation grant Z210009 (to Yan Y.); National Science and Technology Innovation 2030 Major Projects, STI2030-Major Projects grant 2022ZD0204800 (to Yan Y.); National Key R&D Program of China grant 2022YFB4700101 (to H.W.); National Natural Science Foundation of China grant 32070987 and 31722025 (to Yan Y.); Chinese Academy of Sciences Key Program of Frontier Sciences grant QYZDB-SSW-SMC019 (to Yan Y.). S.L. is funded as Chairholder from the Canada Excellence Research Chair in Neuroplasticity; as Research Director at the Belgian National Fund for Scientific Research; the European Foundation of Biomedical Research, the Foundation for Research and Rehabilitation of Neurodegenerative Diseases and the National Natural Science Foundation of China (Grant No.: 81920108023).

We thank members of our laboratory for helpful comments on an earlier version of the manuscript and for helpful discussions. Fred Yang Qu for suggestions on our figure illustration on an earlier version. We thank the License of Krauth/Morel thalamic atlas provided by Axel Krauth, Rémi Blanc, Alejandra Poveda, Daniel Jeanmonod, Anne Morel, and Gábor Székely at University of Zurich and ETH Zurich.

## Author contributions

Conceptualization: Yan Y. and H.W.;

Methodology: Yan Y. and H.W.;

Investigation: H.W., Yan Y., Y.H., P.Z., S.H., S.L., Y.D., Yi Y., Q.G., L.X., X.X., and J.H.;

Clinical Data Resources: Y.D., Yi Y., Q.G., L.X., X.X., and J.H.;

Formal Analysis: H.W. and Y.H.;

Data Curation: H.W., Yan Y. and J.H.;

Visualization: H.W., Yan Y., Y.H. and S.H.;

Funding Acquisition: Yan Y., H.W. and S.L.;

Project Administration: Yan Y.;

Supervision: Yan Y.;

Writing – Original Draft: Yan Y. and H.W.;

Writing – Review & Editing: Yan Y., H.W., Y.H., P.Z., S.H., S.L., and J.H.

---

**Extended Data Fig. 1.**
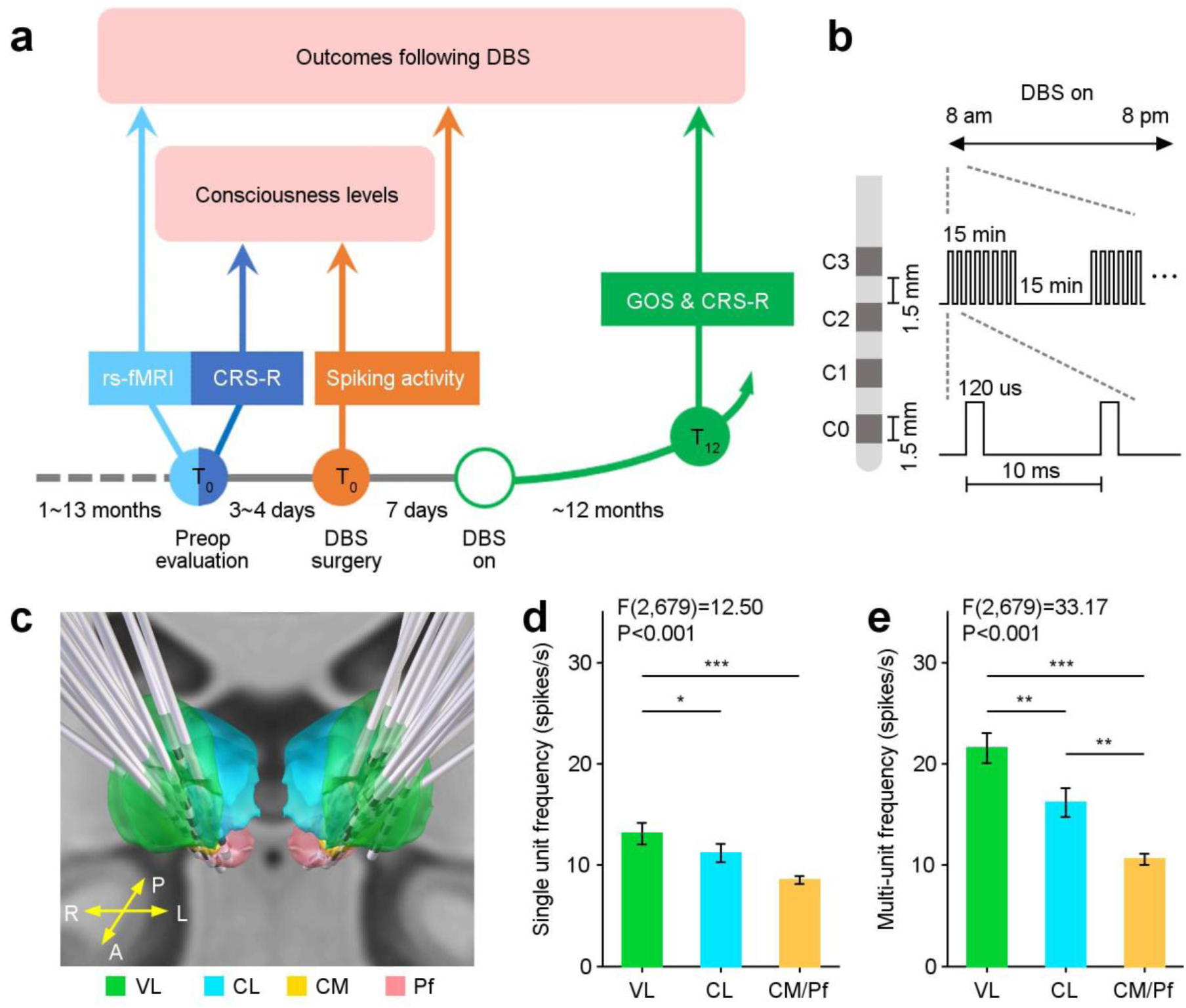
Timeline illustrating the procedure throughout different phases of the study. **a**, For each patient, CRS-R scores and rs-fMRI were acquired three to four days prior to surgery. Thalamic activity was recorded during DBS surgery. These data were collected around the time of the operation, or T0. After seven days of recuperation, DBS was applied. The GOS were utilized to assess the recovery outcomes within 12 months (referred to as T12). The follow-up CRS-R scores were also assessed to monitor the consciousness states following DBS. In the study, we investigated the correlations between neuronal signatures and preoperative consciousness states using spiking activity. By analyzing spiking activities, rs-fMRI and GOS scores, we also evaluated associations between neuronal signatures, brain connections and recovery outcomes following DBS. **b**, The DBS parameters applied to DoC patients. **c**, Reconstructed trajectories of DBS leads for the 29 patients. **d**, **e**, The single-unit frequency and the multi-unit frequency in the three thalamic nuclei according to reconstructed trajectories. The thalamic neuronal firing rates decreased upon the introduction of microelectrodes through the VL, CL to CM/Pf. Two-tailed *P* values for multiple comparisons analysis following ANOVA were corrected using the Tukey-Kramer method. *, *P* < 0.05; **, *P* < 0.01; ***, *P* < 0.001. Error bars: ±SEM.

**Extended Data Fig. 2.**
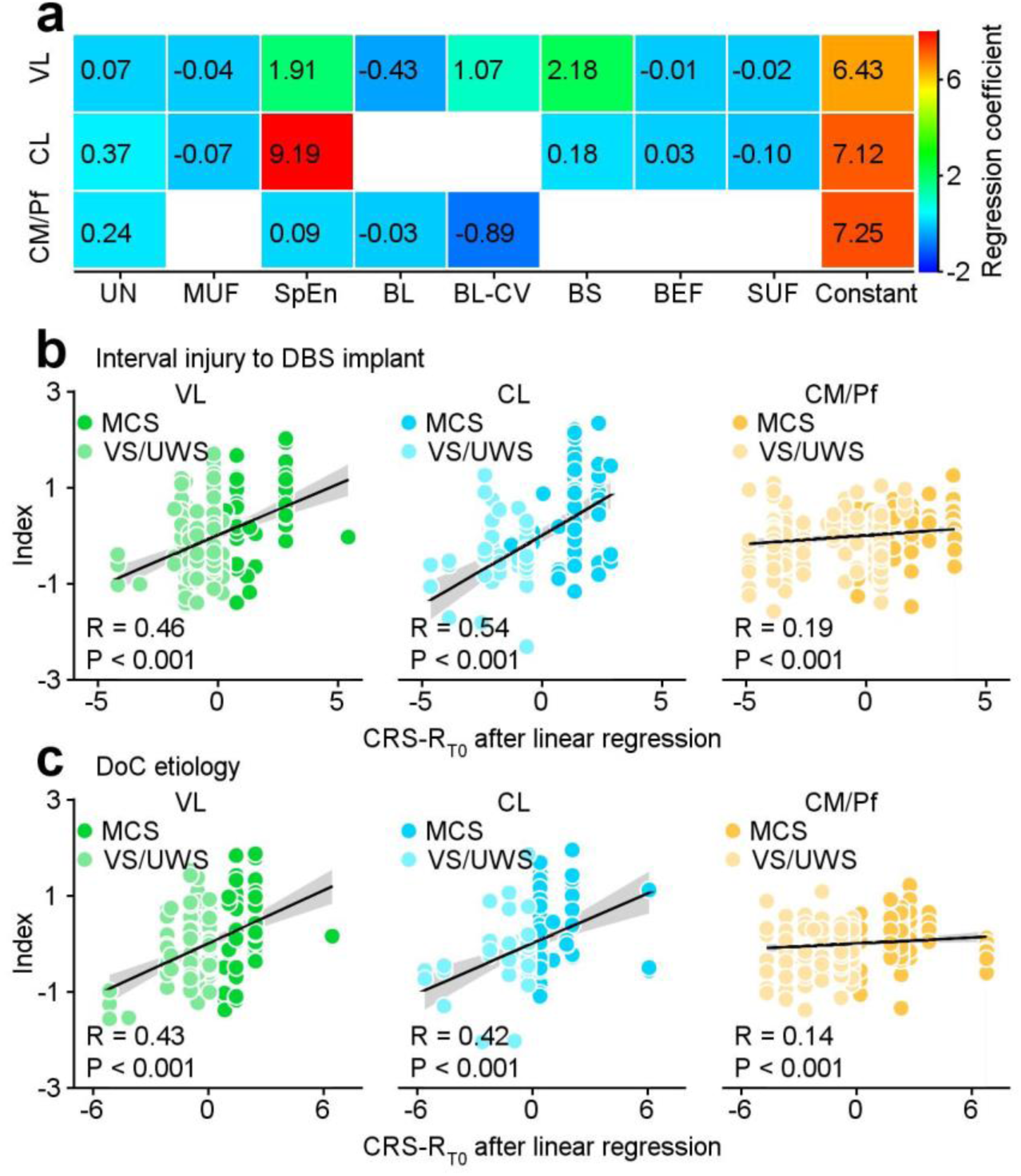
The neuronal index, as defined by PLS regression coefficients, and its relationships with interval injury and etiology. **a**, The neuronal index was determined based on the obtained PLS regression coefficients. The matrix represents the values of the constant and regression coefficients for three thalamic nuclei. **b**, **c**, A linear regression was employed to eliminate the effects of the interval injury to DBS implant (**b**), and etiology (**c**) on neuronal signatures in three thalamic nuclei for all 29 patients. Subsequently, the neuronal indices were re-computed after the liner regression and calculated the Pearson Correlation Coefficient. Linearly fitted lines (black) and its 95% confidence interval (gray) were shown.

**Extended Data Fig. 3.**
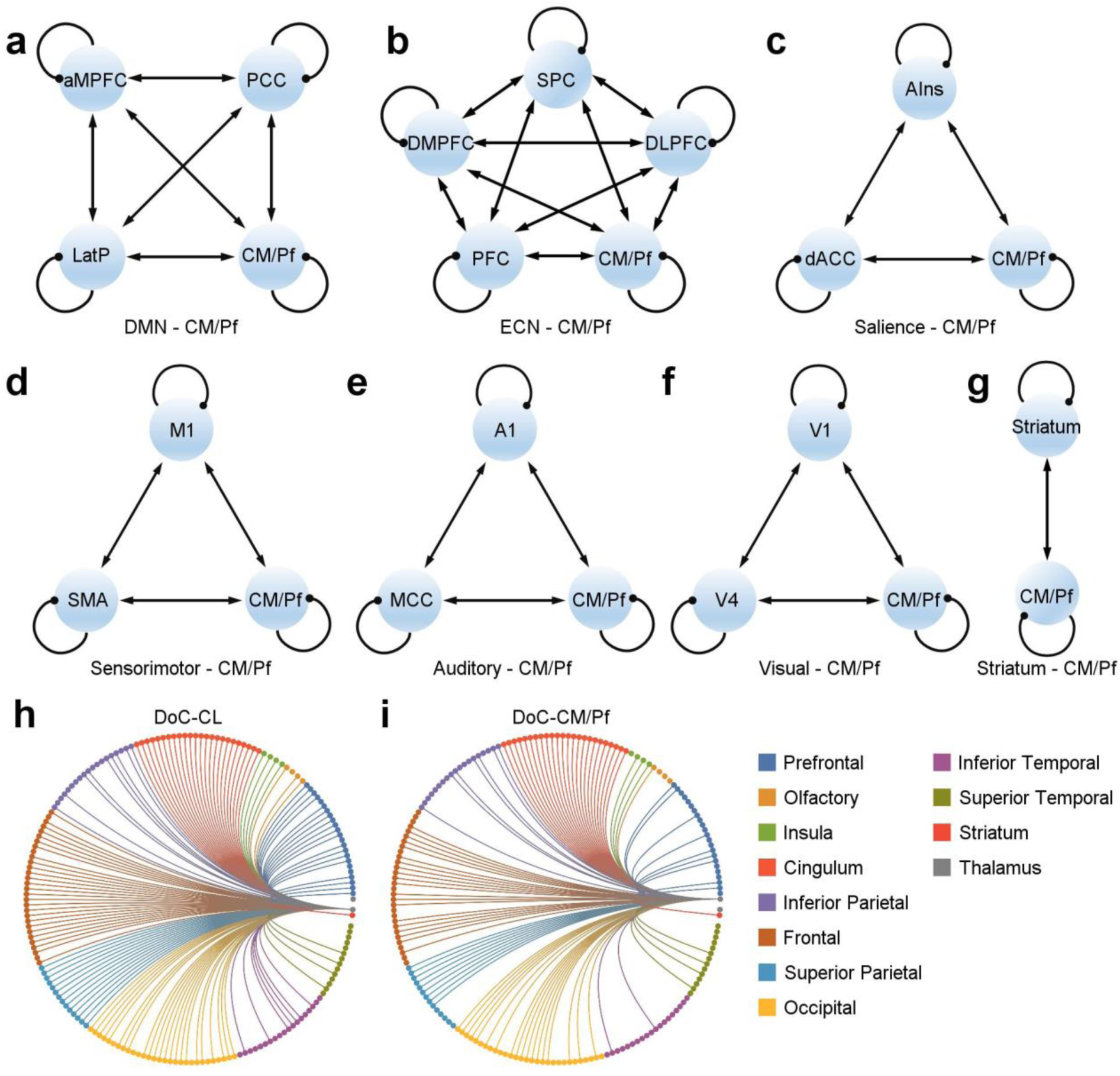
Quantifying effective connectivity of thalamocortical and thalamostriatal connections by utilizing DCM models. **a-f**, Six thalamocortical DCM models featured CM/Pf and the brain regions in a given network (**a**: default mode network; **b**: executive control network; **c**: salience network; **d**: sensorimotor network; **e**: auditory network; **f**: visual network). The effective connectivity (arrows) includes the reciprocal connections between these brain regions, and their self-connections within each region. **g**, The DCM model for the thalamostriatal connection involving the CM/Pf and striatum. **h**, **i**, The structural connectome of the CL (**h**) and the CM/Pf (**i**) in DoC patients.

**Extended Data Fig. 4.**
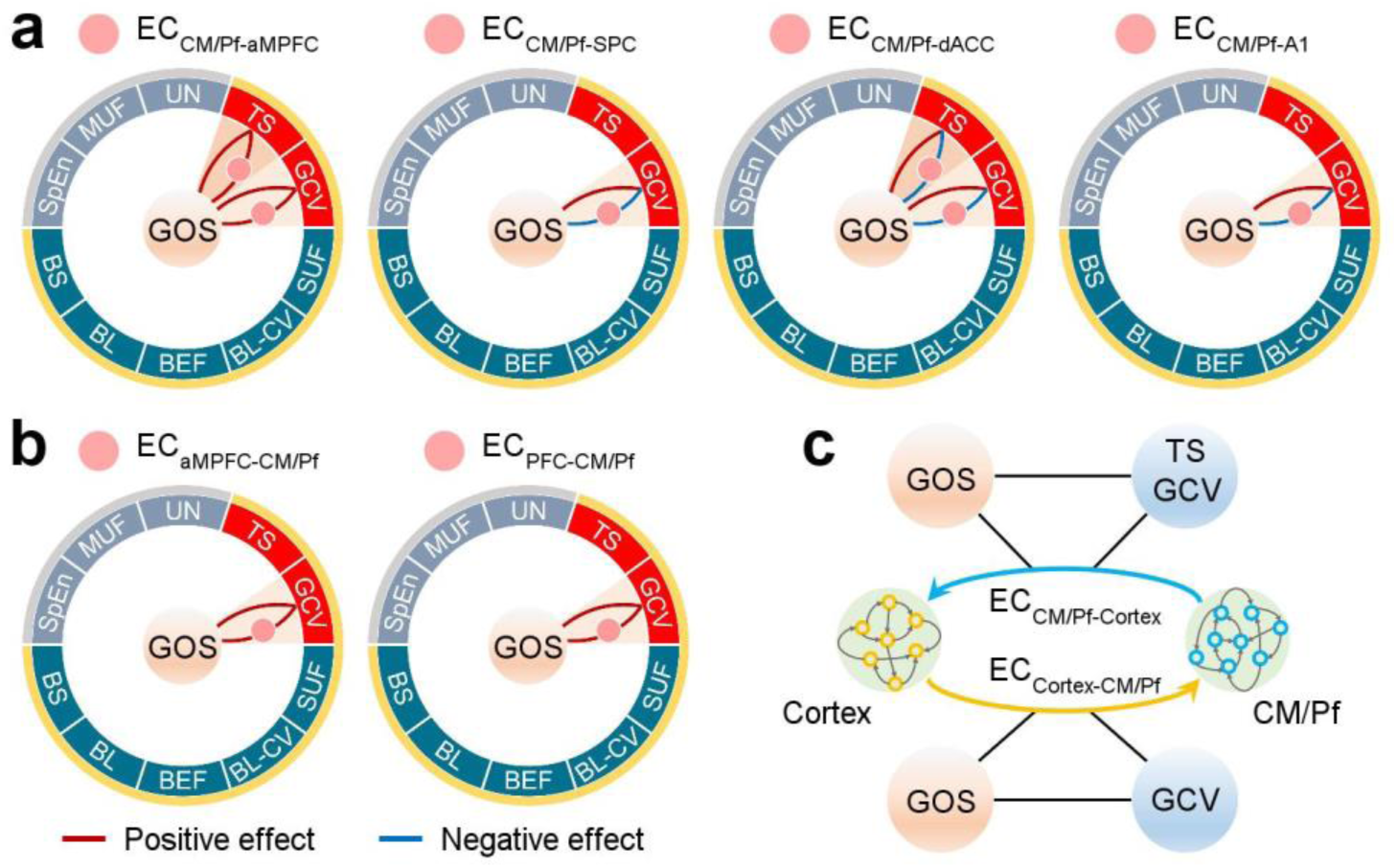
Bidirectional effective connections between CM/Pf and cortical areas formed “logic loops” with neuronal signatures and GOS scores. **a**, Logic loops involved four ECs from the CM/Pf to the cortex. These four cortical areas included anterior medial prefrontal cortex (aMPFC), superior parietal cortex (SPC), dorsal anterior cingulate cortex (dACC), and primary auditory cortex (A1). **b**, Logic loops involved two ECs from the cortex to the CM/Pf, with the two cortical areas, the aMPFC and the anterior prefrontal cortex (PFC). **c**, Summary of logic loops concerning bidirectional ECs, TS and GCV, and GOS scores in (**a**) and (**b**). The tonic firing in the CM/Pf is specifically associated to the unidirectional connections from the CM/Pf to the cortex (blue arrow), rather than the connections in the opposite direction (yellow arrow).

**Extended Data Fig. 5.**
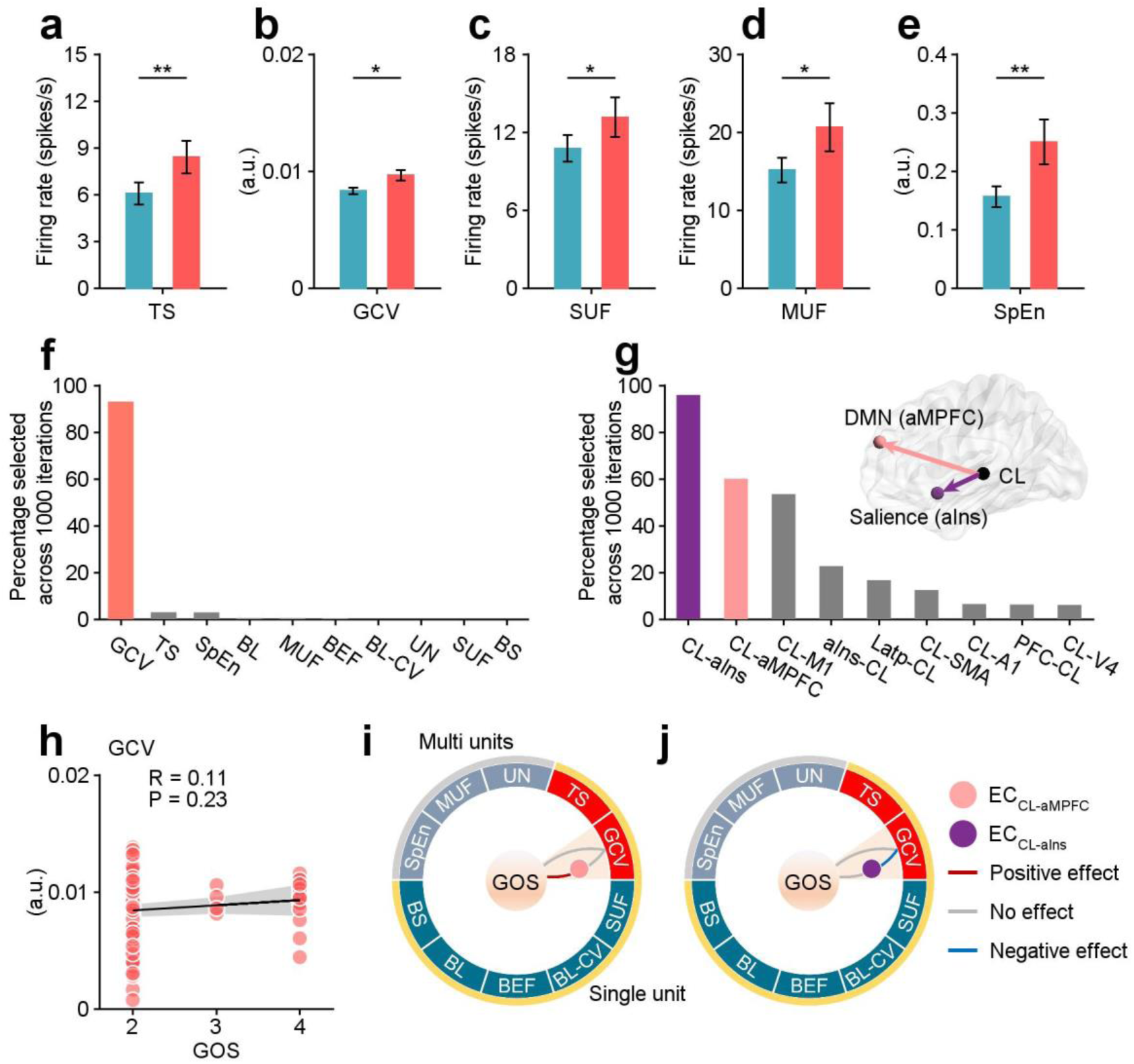
The relationships among neuronal signatures in the CL, thalamocortical connections and recovery outcomes. **a-e**, Neuronal signatures in the CL showed differences between CR (23 recording sites) and CNR (99 recording sites) groups (**a**: tonic spikes; **b**: geometric coefficient of variation; **c**: single-unit frequency; **d**: multi-unit frequency; **e**: sample entropy). *, *P* < 0.05; **, *P* < 0.01; Mann-Whitney-Wilcoxon test. error bars: ± SEM. **f**, The feature selection of neuronal signatures in the CL to discriminate the CR and CNR groups, following a liner regression analysis on the impact of different states of consciousness. **g**, The feature selection of ECs between the CL and cortical areas, as well as the striatum to discriminate the CR and CNR groups. The ECCL-aMPFC and ECCL-aIns were selected based on F scores of ANOVA test. **h**, The Pearson correlation between GCV and GOS scores with *P* = 0.23. Linearly fitted line (black) and its 95% confidence interval (gray shading) were shown. **i**, **j**, The GCV in the CL did not show a significant relationship with either GOS scores or ECs. Therefore, the logic loops are absent for both ECCL-aMPFC (**i**) and ECCL-aIns (**j**).

**Extended Data Table 1.**
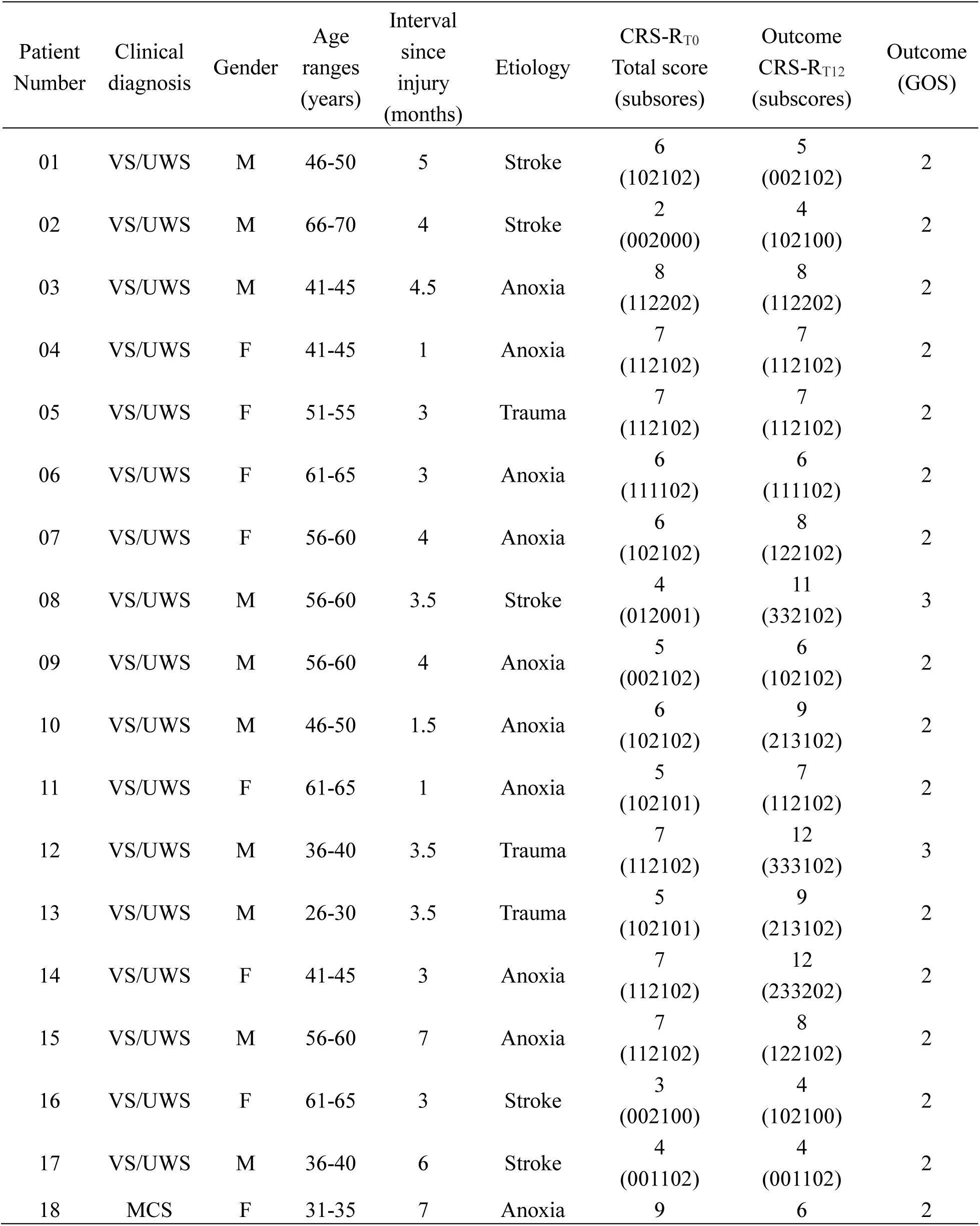
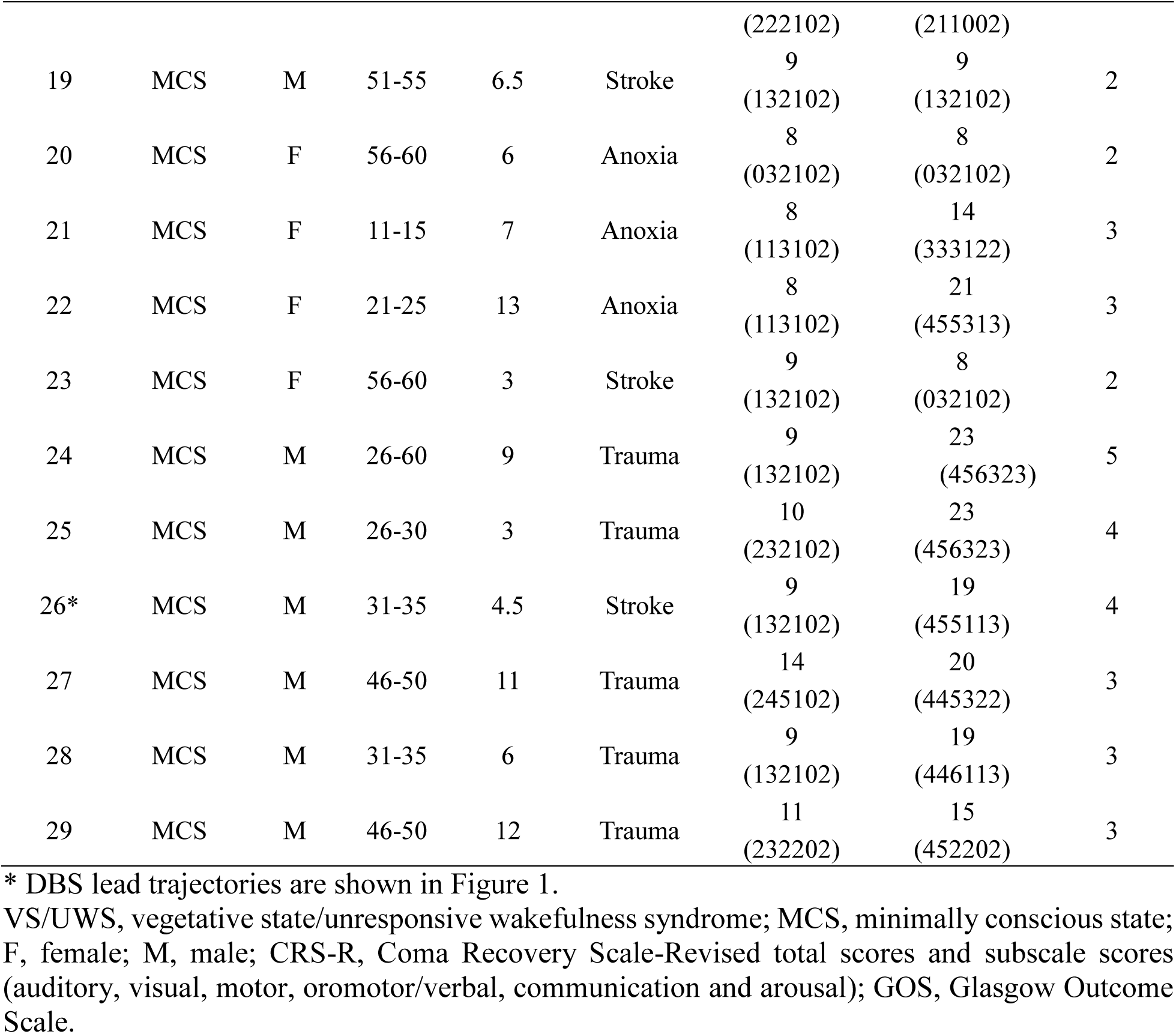
Patient demographic data, etiology, clinical scores and outcome measures.

**Extended Data Table 2.** Acquisition parameters for patients with resting state functional MRI (rs-fMRI) and diffusion MRI data.

| No. | MRI Scanner | rs-fMRI |  |  |  |  |  |  |  | Diffusion MRI |  |  |  |  |  |  |  |  |
| --- | --- | --- | --- | --- | --- | --- | --- | --- | --- | --- | --- | --- | --- | --- | --- | --- | --- | --- |
|  |  | TR<br>(ms) | TE<br>(ms) | Flip<br>angle<br>(°) | FOV<br>(mm) | Matrix | Slices | Voxel size<br>(mm<sup>3</sup>) | Volumes | TR<br>(ms) | TE<br>(ms) | Flip<br>angle<br>(°) | b-values<br>(s/mm<sup>2</sup>) | Directions | FOV<br>(mm) | Matrix | Slices | Voxel size<br>(mm<sup>3</sup>) |
| 01 | GE signa<br>HDxt |  |  |  |  |  |  |  |  | 8000 | 87.4 | 90 | 1000 | 15 | 240 ×<br>240 | 256 ×<br>256 | 30 | 0.94 ×<br>0.94 × 5 |
| 03 | GE signa<br>HDxt |  |  |  |  |  |  |  |  | 17000 | 86 | 90 | 1000 | 32 | 256 ×<br>256 | 256 ×<br>256 | 66 | 1 × 1 × 2 |
| 04 | GE discovery<br>MR750 | 2000 | 30 | 90 | 240 ×<br>240 | 64 ×<br>64 | 39 | 3.75 ×<br>3.75 × 4 | 210 | 9000 | 80.8 | 90 | 1000 | 32 | 256 ×<br>256 | 128 ×<br>128 | 75 | 2 × 2 × 2 |
| 05 | GE discovery<br>MR750 | 2000 | 30 | 90 | 240 ×<br>240 | 64 ×<br>64 | 39 | 3.75 ×<br>3.75 × 4 | 210 | 9000 | 80.8 | 90 | 1000 | 32 | 256 ×<br>256 | 128 ×<br>128 | 75 | 2 × 2 × 2 |
| 07 | GE discovery<br>MR750 | 2000 | 30 | 90 | 240 ×<br>240 | 64 ×<br>64 | 39 | 3.75 ×<br>3.75 × 4 | 210 | 9000 | 80.8 | 90 | 1000 | 64 | 256 ×<br>256 | 128 ×<br>128 | 75 | 2 × 2 × 2 |
| 08# | GE signa<br>HDxt | 2000 | 30 | 90 | 210 ×<br>210 | 64 ×<br>64 | 32 | 3.28 ×<br>3.28 × 4 | 210 | 17000 | 85.4 | 90 | 1000 | 32 | 256 ×<br>256 | 256 ×<br>256 | 66 | 1 × 1 × 2 |
| 09 | GE discovery<br>MR750 |  |  |  |  |  |  |  |  | 9000 | 81.4 | 90 | 1000 | 64 | 256 ×<br>256 | 256 ×<br>256 | 75 | 1 × 1 × 2 |
| 10 | GE discovery<br>MR750 | 2000 | 30 | 90 | 240 ×<br>240 | 64 ×<br>64 | 39 | 3.75 ×<br>3.75 × 4 | 210 | 9000 | 81 | 90 | 1000 | 64 | 256 ×<br>256 | 256 ×<br>256 | 75 | 1 × 1 × 2 |
| 11 | GE signa<br>HDxt | 2000 | 30 | 90 | 192 ×<br>192 | 64 ×<br>64 | 32 | 3 × 3 × 4 | 210 | 17000 | 86 | 90 | 1000 | 32 | 256 ×<br>256 | 256 ×<br>256 | 66 | 1 × 1 × 2 |
| 12 | GE signa<br>HDxt | 2000 | 30 | 90 | 210 ×<br>210 | 64 ×<br>64 | 32 | 3.28 ×<br>3.28 × 4 | 210 | 17000 | 85.4 | 90 | 1000 | 32 | 256 ×<br>256 | 256 ×<br>256 | 66 | 1 × 1 × 2 |
| 13 | GE signa<br>HDxt | 2000 | 30 | 90 | 220 ×<br>220 | 64 ×<br>64 | 32 | 3.44 ×<br>3.44 × 4 | 210 | 17000 | 85.5 | 90 | 1000 | 32 | 256 ×<br>256 | 256 ×<br>256 | 66 | 1 × 1 × 2 |
| 14 | GE signa<br>HDxt | 2000 | 30 | 90 | 220 ×<br>220 | 64 ×<br>64 | 32 | 3.44 ×<br>3.44 × 4 | 210 | 17000 | 85.4 | 90 | 1000 | 32 | 256 ×<br>256 | 256 ×<br>256 | 66 | 1 × 1 × 2 |
| 15 | SIEMENS<br>Verio | 2000 | 30 | 90 | 220 ×<br>220 | 64 ×<br>64 | 32 | 3.44 ×<br>3.44 ×<br>3.45 | 210 |  |  |  |  |  |  |  |  |  |
| 16 | GE signa<br>HDxt | 2000 | 30 | 90 | 192 ×<br>192 | 64 ×<br>64 | 32 | 3 × 3 × 4 | 210 | 17000 | 85.4 | 90 | 1000 | 32 | 256 ×<br>256 | 256 ×<br>256 | 66 | 1 × 1 × 2 |
| 17 | GE signa<br>HDxt | 2000 | 30 | 90 | 220 ×<br>220 | 64 ×<br>64 | 32 | 3.44 ×<br>3.44 × 4 | 210 | 17000 | 85.4 | 90 | 1000 | 32 | 256 ×<br>256 | 256 ×<br>256 | 66 | 1 × 1 × 2 |
| 18 | GE signa<br>HDxt |  |  |  |  |  |  |  |  | 17000 | 85.5 | 90 | 1000 | 32 | 256 ×<br>256 | 256 ×<br>256 | 66 | 1 × 1 × 2 |
| 19 | GE signa<br>HDxt | 2000 | 30 | 90 | 220 ×<br>220 | 64 ×<br>64 | 32 | 3.44 ×<br>3.44 × 4 | 210 | 17000 | 85.4 | 90 | 1000 | 32 | 256 ×<br>256 | 256 ×<br>256 | 66 | 1 × 1 × 2 |
| 22 | GE signa<br>HDxt |  |  |  |  |  |  |  |  | 17000 | 85.4 | 90 | 1000 | 32 | 256 ×<br>256 | 256 ×<br>256 | 66 | 1 × 1 × 2 |
| 23 | GE signa<br>HDxt | 2000 | 30 | 90 | 220 ×<br>220 | 64 ×<br>64 | 32 | 3.44 ×<br>3.44 × 4 | 210 | 17000 | 85.4 | 90 | 1000 | 32 | 256 ×<br>256 | 256 ×<br>256 | 66 | 1 × 1 × 2 |
| 25 | GE signa<br>HDxt | 2000 | 30 | 90 | 220 ×<br>220 | 64 ×<br>64 | 32 | 3.44 ×<br>3.44 × 4 | 240 | 17000 | 85.4 | 90 | 1000 | 32 | 256 ×<br>256 | 128 ×<br>128 | 66 | 2 × 2 × 2 |
| 26 | GE signa<br>HDxt | 2000 | 30 | 90 | 192 ×<br>192 | 64 ×<br>64 | 32 | 3 × 3 × 4 | 210 | 17000 | 85.4 | 90 | 1000 | 32 | 256 ×<br>256 | 256 ×<br>256 | 66 | 1 × 1 × 2 |
| 27 | GE discovery<br>MR750 | 2000 | 30 | 90 | 240 ×<br>240 | 64 ×<br>64 | 39 | 3.75 ×<br>3.75 × 4 | 210 |  |  |  |  |  |  |  |  |  |
| 28 | GE signa<br>PET/MR | 2000 | 30 | 72 | 220 ×<br>220 |  | 36 | 3.44 ×<br>3.44 × 3.8 | 200 |  |  |  |  |  |  |  |  |  |

TR, repetition time; TE, echo time; FOV, field of view.

